# Analysis of large-language model versus human performance for genetics questions

**DOI:** 10.1101/2023.01.27.23285115

**Authors:** Dat Duong, Benjamin D. Solomon

**Affiliations:** Medical Genomics Unit, Medical Genetics Branch, National Human Genome Research Institute, Bethesda, Maryland, United States of America

## Abstract

Large-language models like ChatGPT have recently received a great deal of attention. To assess ChatGPT in the field of genetics, we compared its performance to human respondents in answering genetics questions (involving 13,636 responses) that had been posted on social media platforms starting in 2021. Overall, ChatGPT did not perform significantly differently than human respondents, but did significantly better on memorization-type questions versus critical thinking questions, frequently provided different answers when asked questions multiple times, and provided plausible explanations for both correct and incorrect answers.

## Introduction

Artificial intelligence (AI) applications, including subsets like deep learning (DL) have strong potential in science and medicine, including the field of clinical genetics and genomics.^1-3^ Recently, large-language models (LLMs) like ChatGPT (https://chat.openai.com/chat) have been receiving attention in many venues, including via demonstration of medical knowledge.^4,5^ By way of background, LLMs use a specific type of DL called a transformer. By training the model on a large dataset of text, it learns to predict the next word in a sentence or set of words following a prompt such as a question. Model training involves unsupervised learning, where the models learn to predict the next word in a sentence without explicit labels or annotations.^6^

We aimed to explore how well ChatGPT would perform compared to human respondents in answering questions about genetics.

## Methods

To help evaluate this model related to the field of genetics, including in comparison to human respondents, we asked ChatGPT to answer a series of multiple-choice questions that had been posted on two social media platforms, Twitter and Mastodon, using the following handles (Twitter, @BenjaminSolomo2; Mastodon, @solomonbenjamind@genomic.social). These questions have been posted weekly or biweekly since 2013, with answers and explanations given at the end of each week; 430 questions have been posed to date. For this analysis, we only used the subset of 85 questions posted starting in 2021, as ChatGPT was trained on data prior to this date, and we wanted to avoid the chance that these same questions were used in ChatGPT’s training data. Via the social media polls, these 85 questions have received a total of 13,636 responses as of January 18, 2023. There were fewer respondents on Mastodon (5 questions) versus Twitter (80 questions). Though these questions were answered anonymously through poll functions on the social media platforms, they have been publicly suggested as being useful to physician geneticists, genetic researchers, genetic counselors, and trainees in these fields to help with board and formal examination preparation and to otherwise test knowledge. The followers of these accounts reflect these fields. The questions cover a variety of topics related to genetics and genomics, including general knowledge, clinical genetics and approach to diagnosis and management, molecular genetics and causes of disease, and inheritance patterns and risk calculations. For this analysis, we did not use questions that involved images, such as questions showing a pedigree or a clinical image.

To ask the questions of ChatGPT, we initially uploaded batches of 10-20 questions into ChatGPT at a time. We chose this batch number, as we noticed that ChatGPT would typically answer ∼10-15 questions prior to ceasing to respond further. Each question was accompanied by four possible answers, only one of which was correct. We did not provide any instructions except for telling ChatGPT to pick the single best answer to each question. We had previously noticed that ChatGPT sometimes provides different answers to some questions when asked multiple times, even when not prompted, so after initially asking the questions, we asked all the questions again. In doing this, we did not provide any feedback to ChatGPT in terms of pointing out which questions were right or wrong, since ChatGPT may modify answers according to prompts. Finally, for questions where ChatGPT answered the question incorrectly on either or both attempts, we asked the question again (a third time), and also asked ChatGPT to provide an explanation for the answer it chose in this final attempt.

Because the sample sizes for some categories were small, results were compared using Fisher’s exact test for count data; two-tailed p values are provided.

## Results

Questions, answers, and explanations (the same materials provided via social media to human respondents), as well as ChatGPT’s explanations for any question that was incorrectly answered by ChatGPT, are provided in Supplemental File 1. Along with other details, a summary of human respondent answers to all of these questions are detailed in Supplementary Table 1; to allow cross-referencing, the questions are numbered according to the same numbering system used on the social media accounts.

**Table 1.** Performance of ChatGPT versus respondents. Unless otherwise noted, calculations here and below were done according to ChatGPT’s initial answers, when the questions were first posed.

|  |  |  |  |
| --- | --- | --- | --- |
| **Total** |  |  |  |
|  | **Correct** | **Incorrect** | **Accuracy** |
| Total |  |  |  |
| ChatGPT | 58 | 27 | 68.2% |
| Respondents | 9448 | 4188 | 69.3% |
| **Most common response** |  |  |  |
|  | **Correct** | **Incorrect** | **Accuracy** |
| ChatGPT | 58 | 27 | 68.2% |
| Respondents | 78 | 7 | 91.8% |

When examining answers from all respondents versus ChatGPT’s initial responses, there was not a statistically significant difference (p = 0.8145). However, if we measured the respondents’ group accuracy by choosing the most commonly selected response as the overall group answer, the respondents significantly outperformed ChatGPT (p = 0.00019). See Table 1.

We were interested in how well ChatGPT performed for questions that were considered to focus on memorization (M) or “fact look-up” versus critical thinking (C). To assess this, we divided the questions into these categories based on our subjective assessment of the question (see Supplementary file 1). ChatGPT performed significantly better for memorization than critical thinking questions (p = 2.344e-05). When comparing the ChatGPT results to human respondent results (Table 2), ChatGPT did not perform significantly differently than the human respondents for memorization questions (p = 0.2635) or critical thinking questions (p = 0.06513).

**Table 2.** Comparison of results of memorization versus critical thinking questions.

|  |  |  |  |
| --- | --- | --- | --- |
| **Memorization** |  |  |  |
|  | **Correct** | **Incorrect** | **Accuracy** |
| ChatGPT | 53 | 13 | 80.3% |
| Respondents | 7132 | 2508 | 74.0% |
| **Critical thinking** |  |  |  |
|  | **Correct** | **Incorrect** | **Accuracy** |
| ChatGPT | 5 | 14 | 26.3% |
| Respondents | 1943 | 2053 | 48.6% |

ChatGPT provided different answers to the same questions frequently, with 13 initial answer changes (15% of the 85 questions). ChatGPT initially gave correct answers 58 times. Of the 58 questions were initially answered correctly, ChatGPT gave the wrong answer for 2 questions (3.4% of these 58 questions) when asked the second time. ChatGPT initially gave incorrect answers 27 times. When the questions were posed again, ChatGPT gave different answers 11 times to these 27 questions (40.7% of these 27 questions). For 8 (29.6%) of these 27 initially incorrect answers, the second set of answers were correct. When asked for explanations about the initially incorrect answers, ChatGPT would again sometimes provide different answers along with the explanation (see details in Supplementary Table 1 and Supplementary File 1). We also noted that ChatGPT sometimes embellished the answer. For example, as shown in the supplemental files, it added the acronym “(ECG)” after correctly answering “Electrocardiogram” once and added the phrase “and segregation testing” after correctly answering “Parental testing” to another question. Both are logically correct but were not part of the answer choices. ChatGPT would also sometimes provide full explanations without being prompted.

ChatGPT’s explanations of wrong answers were all relatively plausible in terms of providing believable, logistically consistent (though sometimes incorrect) explanations. Of the explanations given when ChatGPT initially provided the wrong answers, ChatGPT subsequently gave the right answer along with the explanation in 10 instances (37.0% of the initial 27 incorrect answers). ChatGPT gave the correct explanation (explaining why the right answer was correct) but still indicated it chose the wrong answer in 2 instances (7.4% of the initial 27 incorrect answers). These were both after previously giving incorrect answers. In 7 instances (25.9% of the initial 27 incorrect answers), ChatGPT appeared to use incorrect information about a particular condition or topic to select the answer; these was frequent related to particularly esoteric subjects. ChatGPT seemed to frequently misinterpret questions involving calculations and inheritance, with 6 incorrect answers (22.2% of the initial 27 incorrect answers). For 2 questions (7.4% of the initial 27 incorrect answers), ChatGPT appeared to misunderstand the question. For the 2 questions that ChatGPT answered correctly initially and then incorrectly the second time, it provided the correct answer and explanation when asked to provide the explanation.

## Discussion

We were impressed with ChatGPT’s performance, including because its answers were almost instantaneous, and were compared to humans who were able to look up answers and discuss the questions through the social media posts (we note that we do not know to what extent respondents selected their answers without looking up information or asking others). ChatGPT performed better with memorization-type questions versus critical thinking-type questions. This aligns with the consensus about its performance, where the assumption is that AI is not yet fully better than humans at critical thinking (for example, as a response to the proliferation of ChatGPT in educational settings, some teachers are redesigning assignments to emphasize critical thinking). This may be a short-term fix; with the ingestion of more data, ChatGPT may bridge this gap soon.

We did not fine-tune the model, which would be expected to improve performance. We note that it is not currently possible to know when ChatGPT is correct or not without additional verification – ChatGPT will offer a plausible explanation for an incorrect answer and may at various times offer completely different responses and explanations. In medical or other high-stakes settings, this is concerning. However, strategies like labeling responses with confidence scores, such as is available in AlphaFold, may help.^7^

Overall, approaches using LLMs have shown that they can perform relatively well in a variety of medical testing situations.^4,5^ These scores are sure to improve, perhaps rapidly, and tools like ChatGPT may become ubiquitous, much the way various forms of AI are currently used in clinical and research genomic analyses every day.^8,9^ As one of many examples, patients and research subjects may increasingly turn to ChatGPT and other such interfaces to learn about conditions that may affect them, including the suggested work-up and treatment. This may increase difficulties with issues like misinformation.

It seems fitting to offer ChatGPT the final words here; after describing the premise of this paper to ChatGPT, we asked it to provide a concluding paragraph (query given 1/18/23 via the link in the Introduction). It wrote the following:

In conclusion, models like ChatGPT have the potential to greatly impact the field of clinical genetics by providing rapid and accurate responses to a wide range of genetic-related questions. This can aid healthcare professionals in the diagnosis and treatment of genetic disorders, as well as provide patients and their families with easily accessible information about their conditions. Additionally, ChatGPT’s ability to understand and respond to natural language queries could make genetic information more widely available to a non-expert audience. As the field of genetics continues to advance, the use of natural language processing models like ChatGPT will become increasingly important in both research and clinical settings.

## Data Availability

All data produced in the present work are contained in the manuscript and supplementary files.

## Data Availability

All data used and presented are available in the manuscript and supporting (supplementary) files.

## Acknowledgments

This research was supported by the Intramural Research Program of the National Human Genome Research Institute, National Institutes of Health.

## Author Contributions

All authors contributed to the conception and design of the work and to acquisition, analysis, and interpretation of data, and both drafted or substantially revised the work.

