## Supplementary File 1 for "Analysis of large-language model versus human performance for genetics questions"

**Supplemental file 1**: Questions, answers and explanations.

Questions were originally posed in week-long Twitter and/or Mastodon polls, using the following handles: Twitter, @BenjaminSolomo2; Mastodon, @solomonbenjamind@genomic.social. Of over 400 questions posed to date, only questions starting in 2021 (the date after which ChatGPT stopped gathering data) are shown. Questions are shown in the same format as in the social media polls, including the individual components (questions, answers, and explanations). Questions that include visual images, such as a pedigree or radiologic image used as part of the question, were excluded from this list. Each question was asked twice. For questions that were answered incorrectly by ChatGPT, ChatGPT’s answers are given for the two different attempts. When ChatGPT gave incorrect answers, the question was repeated again, and ChatGPT was asked to provide an explanation, which is appended.

Q325

Acrodermatitis enteropathica, Birk-Landau-Perez syndrome, and Spondylocheirodysplastic Ehlers-Danlos syndrome are genetic conditions that all involve abnormalities related to metabolism of what metal?

Copper

Iron

Magnesium

Zinc

Q325 answer: Zinc

<https://pubmed.ncbi.nlm.nih.gov/29354481/>

Q325 explanation

Multiple genetic disorders are due to abnormalities related to the metabolism of different metals, including copper, iron, and zinc. Of these, Acrodermatitis enteropathica, Birk-Landau-Perez syndrome, and Spondylocheirodysplastic Ehlers-Danlos syndrome, as well as Transient neonatal zinc deficiency, which represent diverse clinical conditions, all involve abnormal zinc metabolism.

Reference(s): Ferreira and Gahl, Transl Sci Rare Dis 2017

ChatGPT first answer: Copper

ChatGPT second answer: Zinc

ChatGPT explanation (when asked the question and to provide an explanation, the correct answer was given):

Zinc. Acrodermatitis enteropathica, Birk-Landau-Perez syndrome, and Spondylocheirodysplastic Ehlers-Danlos syndrome are all genetic conditions that involve abnormalities related to the metabolism of zinc. Acrodermatitis enteropathica is a rare inherited disorder characterized by a deficiency of zinc in the body. Birk-Landau-Perez syndrome is a rare genetic disorder characterized by zinc deficiency. Spondylocheirodysplastic Ehlers-Danlos syndrome is a rare genetic disorder characterized by abnormal connective tissue, caused by a deficiency of zinc in the body. In all these conditions, the body is unable to absorb or utilize zinc properly, leading to a deficiency of this essential mineral.

Q326

Which of the following would represent a frameshift variant?

c.169_170insA

c.79_80delinsTT

g.123A>G

p.Trp24Cys

Q326 answer: c.169_170insA

<https://www.cancer.gov/publications/dictionaries/genetics-dictionary/def/frameshift-variant>

<https://www.ncbi.nlm.nih.gov/pubmed/27367167>

<http://varnomen.hgvs.org/>

Q326 explanation

A frameshift variant involves the insertion or deletion of a number of base pairs that is not a multiple of three. This change alters the DNA sequence’s triplet reading frame, and usually results in a truncated protein product due to a premature stop codon.

Reference(s): den Dunnen, Curr Protoc Hum Genet 2016

Q327

Approximately what percentage of the human genome consists of repetitive elements?

10%

25%

50%

99%

Q327 answer: 50%

<https://pubmed.ncbi.nlm.nih.gov/17187668/>

<https://pubmed.ncbi.nlm.nih.gov/29398703/>

<https://pubmed.ncbi.nlm.nih.gov/33442040/>

Q327 explanation

Repetitive elements are found frequently in the genomes of both prokaryotes and eukaryotes. It is estimated that about half of the human genome consists of repetitive elements (this is sometimes called the “repeatome”).  However, estimates do vary considerably.  Though challenging to accurately sequence and analyze, there is growing evidence that variants in these repetitive elements can contribute to a number of different human diseases, including in complex/multifactorial ways.

Reference(s): Haubold and Wiehle, BMC Bioinformatics 2006; Tannan, Nat Rev Genet 2018; Mitra et al., Nature 2021

Q328

On examination, a patient is noted to have “coarse” facial features.  Based on this finding, which type of testing would most likely be indicated?

Chromosomal breakage

FISH for 22q11.2

FMR1 repeat analysis

Urine glycosaminoglycans

Q328 answer: Urine glycosaminoglycans

<https://elementsofmorphology.nih.gov/index.cgi?tid=39ff7c97ca920a5d>

<https://pubmed.ncbi.nlm.nih.gov/23958290/>

<https://pubmed.ncbi.nlm.nih.gov/32235807/>

Q328 explanation

“Coarse” facial features occur due to thickened skin (with or without thickening of the underlying tissues) or more rounded facial features, and generally refers to a less sharp appearance of the facial structures. Coarse facial features occur in a number of different genetic conditions, including mucopolysaccharidoses, of which there are multiple different types. In a patient with a suspected mucopolysaccharidosis the diagnostic process can include testing of urinary glycosaminoglycans (GAGs) as an initial part of the overall work-up. Accurate diagnosis (including via more specific testing modalities than urinary GAGs) can be important to ensure precise diagnosis and medical management of the conditions.

Reference(s): NHGRI <https://elementsofmorphology.nih.gov/>; Lawrence et al., Mol Genet Metab 2014; Kubaski et al., Diagnostics (Basel) 2020

ChatGPT first answer: FMR1 repeat analysis

ChatGPT second answer: FMR1 repeat analysis

ChatGPT explanation (when asked the question and to provide an explanation, the correct answer was given):

FISH for 22q11.2. "Coarse" facial features can be a sign of a genetic disorder known as 22q11.2 deletion syndrome, also known as DiGeorge syndrome or Velocardiofacial syndrome. This disorder is caused by a deletion of a small piece of chromosome 22, specifically at the q11.2 locus, which results in a loss of multiple genes in this region. One of the most common ways to diagnose 22q11.2 deletion syndrome is by using a technique called fluorescence in situ hybridization (FISH), which can detect the deletion on chromosome 22. This is the most likely test that would be indicated based on the physical finding of "coarse" facial features

Q329

A man and a woman have 2 children, both of whom are affected by an autosomal recessive condition. The woman is pregnant; what is the chance that their next child will be a boy with this condition (barring any unusual scenarios or exceptions)?

0.025%

12.5%

25%

33%

Q329 answer: 12.5%

Q329 explanation

In the question above, both parents are presumed to be heterozygous “carriers” for a pathogenic variant in the relevant gene. Except for certain unusual circumstances, the chance for any of their children to be affected is 25%.  This does not depend on the number of previously affected or unaffected children. Again, assuming that there are no unusual circumstances, the chance for a child to be a boy is approximately 50%.  Thus, the chance for the next child to be an affected boy= 25% x 50% = (0.25)(0.5) = 0.125, or 12.5%

Reference(s): Thompson & Thompson, Genetics in Medicine, 7th ed.

ChatGPT first answer: 25%

ChatGPT second answer: 25%

Chat GPT explanation:

25%. Autosomal recessive conditions are inherited when an individual receives two copies of a mutated gene, one from each parent. The parents in this scenario are known carriers of the condition, meaning they each have one copy of the mutated gene but do not show symptoms of the condition.

The chance that their next child will be a boy is 50%. The chance that a child will inherit the mutated gene from both parents, and therefore be affected by the condition, is 25% (1/4). Therefore, the chance that their next child will be a boy with this condition is 25%.

It's worth noting that this probability is based on the assumption that both parents are carriers of the same autosomal recessive condition, and assuming Mendelian inheritance pattern. If the parents are carriers of different mutations in the same gene, the chance of having an affected offspring may vary. And if the parents have a different recessive condition, the chance will change accordingly.

Q330

An infant was slightly premature and is being taken care of in the NICU; on rounds, the team notes “puffiness” under the eyes, and that the calcium level has been high on routine blood chemistries.  What genetic condition might be suspected based on this?

22q11.2 deletion syndrome

Homocystinuria

Marfan syndrome

Williams syndrome

Q330 answer: Williams syndrome

<https://pubmed.ncbi.nlm.nih.gov/12088082/>

<https://www.ncbi.nlm.nih.gov/books/NBK1249/>

<https://pubmed.ncbi.nlm.nih.gov/31964759/>

Q330 explanation

Williams syndrome, which is estimated to occur in more than 1 in 10,000 births, occurs due to heterozygous deletion of chromosome 7q11.23. The condition affects multiple organ systems, and often includes a distinctive facial appearance that evolves as the person ages (in infancy this can include periorbital fullness, as well as multiple other findings), cardiovascular anomalies (eg, supravalvar aortic stenosis), endocrine disorders (including neonatal hypercalcemia and hypercalciuria), and other features. Accurate diagnosis is important to ensure optimal care.

Reference(s): Strømme et al., J Child Neurol 2002; Morris, GeneReviews 2017; Morris et al., Pediatrics 2020

ChatGPT first answer: 22q11.2 syndrome

ChatGPT second answer: 22q11.2 syndrome

ChatGPT explanation (when asked the question and to provide an explanation, the correct answer was given):

Williams syndrome. Williams syndrome is a genetic disorder that is characterized by several physical and developmental features, one of which is puffiness under the eyes. The medical term for puffiness under the eyes is "epicanthal folds" and is one of the diagnostic criteria for Williams Syndrome. Additionally, individuals with Williams syndrome have elevated calcium levels on routine blood chemistries, which is caused by hypercalcemia. The combination of "puffiness" under the eyes and high calcium levels on routine blood chemistries would be considered classic features of Williams syndrome. It is a rare genetic disorder caused by the deletion of genetic material from a specific region of chromosome 7.

Q331

A patient has features consistent with Turner syndrome. Assuming this is the correct diagnosis, what result would be expected on routine cytogenomic studies (with the caveat that mosaic results are not uncommon)?

45,X

45,XO

46,XX

47,XXX

Q331 answer: 45,X

<https://pubmed.ncbi.nlm.nih.gov/11498234/>

<https://pubmed.ncbi.nlm.nih.gov/30790455/>

<https://pubmed.ncbi.nlm.nih.gov/30790449/>

Q331 explanation

Turner syndrome is a genetic condition that occurs in about 1 in 2,500 live-born female infants (though estimates vary). Turner syndrome is due to absence of all or part of  the X chromosome. About half of people with Turner syndrome have monosomy X (which is written as 45,X, not 45,XO). Approximately 10% of individuals with Turner syndrome have a duplication (isochromosome) of the long arm of one of the X chromosomes. Most of the rest of people with Turner syndrome will have mosaicism for 45,X along with the presence of additional cell lineages, such as 46,XX.

Reference(s): Ranke and Saenger, Lancet 2001; Kruszka and Silberbach, Amer J Med Genet C 2019; San Roman and Page, Amer J Med Genet C 2019

Q332

Specific genetic changes from genetic testing can help guide therapy. In a patient with certain types of cancer, identification of a germline pathogenic variant in BRCA1 or BRCA2 might lead to consideration of therapy with what type of agent?

AAV9-based gene therapy

Nitrosourea

PARP inhibitor

PD-1 inhibitor

Q332: PARP inhibitor

<https://pubmed.ncbi.nlm.nih.gov/14576434/>

<https://www.ncbi.nlm.nih.gov/books/NBK1247/>

<https://pubmed.ncbi.nlm.nih.gov/30110579/>

<https://pubmed.ncbi.nlm.nih.gov/31085765/>

<https://pubmed.ncbi.nlm.nih.gov/32493233/>

Q332 explanation

Pathogenic variants (mutations) in the BRCA1 or BRCA2 genes can be involved in an increased risk of a number of cancer types, including breast cancer (in both males and females), ovarian cancer, and prostate cancer. Variants in specific genes may help drive the selection of therapies as well as the overall management strategy for many types of cancer; this may be true of variants found in the tumors (often called “somatic variants” in this context) or germline variants. Clinical trials have shown efficacy of PARP (poly-ADP ribose polymerase) inhibitors (e.g., olaparib, niraparib, and rucaparib) for individuals with certain cancer types related to germline pathogenic variants in BRCA1 and BRCA2, and drugs in this class have been approved in certain scenarios.

Reference(s): King et al., Science 2003; Petrucelli et al., GeneReviews 2016; Litton et al., N Engl J Med 2018; Cheng et al., J Natl Compr Canc Netw 2019; Rehman Mohyuddin et al., BMC Cancer 2020

Q333

Which of the following is a membrane-bound organelle that contains digestive enzymes and is involved in a number of cellular processes, including apoptosis, metabolism of large molecules made by the body, and destruction of pathogens?

Golgi body

Lysosome

Mitochondrion

Ribosome

Q333: Lysosome

<https://www.genome.gov/genetics-glossary/Lysosome>

<https://pubmed.ncbi.nlm.nih.gov/19336380/>

<https://pubmed.ncbi.nlm.nih.gov/31140601/>

Q333 explanation

Lysosomes are membrane-bound organelles that contain digestive enzymes. Lysosomes are involved in many cellular processes, including the metabolism of large molecules, destruction of pathogens, and apoptosis. A number of genetic conditions are considered lysosomal disorders, and are due to genetic changes that affect specific lysosomal functions. Examples of lysosomal disorders include Batten disease, cystinosis, Fabry disease, Gaucher disease, mucolipidoses, mucopolysaccharide storage diseases, and Pompe disease, to name just a few of the more than 50 different conditions. Accurate diagnosis can be critical to ensure appropriate management, which can significantly impact morbidity and mortality for many conditions.

National Human Genome Research Institute Talking Glossary of Genetic Terms; Staretz-Chacham et al., Pediatrics 2009; Lachmann, J Inherit Metab Dis 2020

Q334

An infant has a positive newborn screen for X-linked adrenoleukodystrophy (X-ALD). Follow-up tests are negative for X-ALD, but the child starts to show signs of developmental delay in infancy. What condition should be considered?

Aarskog syndrome

Aicardi-Goutières syndrome

Alagille syndrome

Alpha thalassemia

Q334 answer: Aicardi-Goutières syndrome

<https://pubmed.ncbi.nlm.nih.gov/17846997/>

<https://pubmed.ncbi.nlm.nih.gov/28739201/>

<https://pubmed.ncbi.nlm.nih.gov/32003821/>

<https://pubmed.ncbi.nlm.nih.gov/33683010/>

Q334 explanation

Newborn screening uses a combination of methods to detect selected, potentially treatable conditions. Screening for X-linked adrenoleukodystrophy (X-ALD) involves assessing for the presence of abnormal levels of C26:0 lysophosphatidylcholine. A number of patients with Aicardi-Goutières Syndrome (AGS) have been identified through newborn screening for X-ALD. AGS can result from pathogenic variants (mutations) in several different genes. AGS is characterized by central nervous system calcifications, leukodystrophy, and neurocognitive impairment. One hint about the possibility of AGS in patients who screen positive for X-ALD involves the presence of early-onset neurological manifestations.

Reference(s): Rice et al., Am J Hum Genet 2007; Armangue et al., Mol Genet Metab 2017; Lee et al., JAMA Netw Open 2020; Tise et al., Am J Med Genet A 2021

Q335

During a viral pandemic, genetic variants that similarly increase transmissibility (and therefore result in a selective advantage) appear independently in different geographic locations. What is the best term for this?

Convergent evolution

Divergent evolution

Mutualism

Parsimony

Q335 answer: Convergent evolution

<https://pubmed.ncbi.nlm.nih.gov/22456349/>

<https://pubmed.ncbi.nlm.nih.gov/27030773/>

<https://pubmed.ncbi.nlm.nih.gov/33594385/>

<https://pubmed.ncbi.nlm.nih.gov/33688681/>

Q335 explanation

Convergent evolution refers to the independent evolution of similar features. One example is the independent occurence of the ability to use wings to fly in very different species; this results in the analogous structure of wings. In the example given in the question, similar variants, all of which increase viral transmissibility, appear in independent virus populations. This is presumably due to selective pressures such that the variant results in some type of selective advantage.

Reference(s): Yang and Rannala, Nat Rev Genet 2012; Chin and Lentink, J Exp Biol 2016; Hodcroft et al., medRxiv 2021; Martin et al., medRxiv 2021

Q336

A patient has undergone extensive genetic testing without finding an explanation. They enroll in a research study involving a new type of genomic analysis. As part of enrollment, informed consent is obtained. What basic ethical principle includes informed consent?

Beneficence

Justice

Respect for persons

Subject selection

Q336 answer: Respect for persons

<https://www.hhs.gov/ohrp/regulations-and-policy/belmont-report/index.html>

<https://www.hhs.gov/ohrp/regulations-and-policy/belmont-report/read-the-belmont-report/index.html>

<https://pubmed.ncbi.nlm.nih.gov/24705284/>

Q336 explanation

The Belmont Report, which derived from formal commissioned work in the 1970s, describes the basic ethical principles that should underlie biomedical and behavioral research involving human subjects. The Belmont Report also developed guidelines based on these principles. The three basic ethical principles described are: Respect for persons; Beneficence; Justice.  Per the report, “respect for persons incorporates at least two ethical convictions: first, that individuals should be treated as autonomous agents, and second, that persons with diminished autonomy are entitled to protection.” Though there are many nuances, including as relates to genomic research, informed consent is considered to be part of Respect for persons.

Reference(s): The Belmont Report (HHS.gov); Mathews and Jamal, Genes (Basel) 2014

Q337

A two year-old girl has been diagnosed with Rett syndrome. In addition to supportive and other care, what type of surveillance is recommended based on this diagnosis?

Colonoscopy

Electrocardiogram

LH and FSH levels

Melanoma screening

Q337 anwer: Electrocardiogram

<https://pubmed.ncbi.nlm.nih.gov/10208957/>

<https://pubmed.ncbi.nlm.nih.gov/28394409/>

<https://www.ncbi.nlm.nih.gov/books/NBK1497/>

Q337 explanation

Rett syndrome can result from pathogenic variants (“mutations”) in the MECP2 gene. Classic Rett syndrome involves females who have apparently normal cognitive development until about 18 months months of life, followed by developmental stagnation, regression, and then long-term stability. The condition is now considered a spectrum of disease that can arise from pathogenic variants affecting the MECP2 gene; similar presentations can involve other genes. One complication of Rett syndrome can include a prolonged corrected QT (QTc) interval, and surveillance via electrocardiogram has been recommended due to this finding, with cardiac follow-up if a prolonged QTc is detected.

Reference(s): Ellaway et al., Arch Dis Child 1999; Crosson et al., Am J Med Genet A 2017; Kaur et al., GeneReviews 2019

Q338

A patient has features consistent with Noonan syndrome. If this is the correct diagnosis, which gene is most likely to be involved?

KRAS

NF1

PTPN11

RAF1

Q338 answer: PTPN11

<https://www.ncbi.nlm.nih.gov/books/NBK1124/>

<https://pubmed.ncbi.nlm.nih.gov/31115195/>

Q338 explanation

Noonan syndrome, like many genetic conditions, can be caused by pathogenic variants (“mutations”) in multiple different genes - this phenomenon is termed genetic heterogeneity.  Among patients with Noonan syndrome, pathogenic variants in the gene PTPN11 are the most frequently identified (in about 50% of individuals). Pathogenic variants in other genes, such as SOS1, RAF1, RIT1, KRAS, BRAF, LZTR1, MAP2K1, and NRAS can also cause the condition, and some genotype-phenotype correlations have been suggested. Variants in these genes may also be related to other conditions.

Reference(s): Allanson and Roberts, GeneReviews 2019; Liao and Mehta, Pediatr Endocrinol Rev 2019

Q339

On physical exam, a child is noted to have unilateral leukocoria (the pupillary reflex appears white during the eye exam). Which condition might be suspected?

Macular degeneration

Retinitis pigmentosa

Retinoblastoma

Tay-Sachs disease

Q339 answer: Retinoblastoma

<https://www.ncbi.nlm.nih.gov/books/NBK1452/>

<https://www.ncbi.nlm.nih.gov/books/NBK560794/>

Q339 explanation

Retinoblastoma, a malignant tumor of the retina, typically occurs in young children, usually before age five years. The condition is related to variants in the gene RB1. Signs of retinoblastoma may include: leukocoria (an abnormally pupillary reflex where there is a white appearance to the pupil on eye examination), strabismus, and decreased vision. Some affected individuals may have other medical or family history suggesting hereditary retinoblastoma.

Reference(s): Lohmann and Gallie, GeneReviews 2019; Kanukollu and Tripathy, StatPearls 2021

Q342

The first form of gene therapy to be FDA-approved for use in the United States, treats what condition?

Acute lymphoblastic leukemia

Cystic fibrosis

Leber congenital amaurosis

Sickle cell anemia

Q342 answer: Acute lymphoblastic leukemia

<https://www.cancer.gov/publications/dictionaries/cancer-terms/def/tisagenlecleucel>

<https://pubmed.ncbi.nlm.nih.gov/25317870/>

<https://pubmed.ncbi.nlm.nih.gov/29385370/>

Q342 explanation

Tisagenlecleucel, a form of gene therapy to treat B-cell acute lymphoblastic leukemia (ALL), was approved by the FDA in 2017. In this approach, T cells are removed from the patient, and are genetically modified to create a chimeric receptor on the surface of the T cells.  That is, a gene to make a chimeric antigen receptor (CAR) is added to the T cells - this is why this type of therapy is sometimes called “CAR-T” therapy.  After this is done in the laboratory, the modified cells are transferred back to the patient. Since the approval of tisagenlecleucel, gene therapies to treat other conditions - including conditions that are more traditionally considered “genetic” or inherited conditions,” such as spinal muscular atrophy and RPE65-associated retinal dystrophy - have also been approved in the US.

Reference(s): Maude et al., N Engl J Med 2014; Maude et al., N Engl J Med 2018

ChatGPT first answer: Leber congenital amaurosis

ChatGPT second answer: Leber congenital amaurosis

ChatGPT explanation:

The first form of gene therapy to be FDA-approved for use in the United States, treats Leber congenital amaurosis. Leber congenital amaurosis (LCA) is a rare inherited eye disease that causes severe vision loss in infants and young children. The first FDA-approved gene therapy, voretigene neparvovec (Luxturna), uses a vector to deliver a normal copy of the RPE65 gene to cells in the retina, allowing them to produce a functional protein that is needed to produce visual pigment. This therapy is intended to help improve vision in people with LCA caused by a specific RPE65 gene mutation.

Q344

An apparently healthy woman gives birth to a term male infant. Her brother had a severe inborn error of metabolism, and testing shows that her newborn son is also affected. Assuming no consanguinity or other such circumstance, what condition is most likely in this family?

Biotinidase deficiency

OTC deficiency

Phenylketonuria

Wilson disease

Q344 answer: OTC deficiency

<https://pubmed.ncbi.nlm.nih.gov/14705115/>

<https://www.ncbi.nlm.nih.gov/books/NBK154378/>

<https://pubmed.ncbi.nlm.nih.gov/27854360/>

Q344 explanation

Ornithine transcarbamylase (OTC) deficiency, caused by pathogenic variants in the OTC gene, is an X-linked urea cycle disorder. In the question, the history of the affected brother, the apparently healthy mother (the sister of the affected brother), and the newborn son with the same condition as his uncle, suggest an X-linked condition. Of note, it is estimated that approximately 20-30% of females harboring pathogenic variants in OTC will display some clinical manifestations. Early diagnosis of affected individuals is important to allow treatment, both for acute episodes, as well as for long-term management.

Reference(s): Gyato et al., Ann Neurol 2004; Lichter-Konecki et al., GeneReviews 2016; Kalia et al., Genet Med 2017

ChatGPT first answer: Phenylketonuria

ChatGPT second answer: Phenylketonuria

ChatGPT explanation:

Phenylketonuria (PKU) is a genetic disorder caused by a deficiency of the enzyme phenylalanine hydroxylase (PAH), which is responsible for converting the amino acid phenylalanine (PHE) to tyrosine. PKU is inherited in an autosomal recessive manner, meaning that an individual must inherit two copies of the mutated gene, one from each parent, in order to develop the condition. It is most likely that the woman is a carrier of PKU, and her newborn son is affected by PKU because the woman's brother had a severe inborn error of metabolism and the newborn son is also affected and both are passing the condition in an autosomal recessive manner. The chance that their next child will inherit the condition would be 25% if the woman's partner is not a carrier.

It's worth noting that PKU is a rare condition, and it's important to confirm the diagnosis with a genetic test, as well as to rule out other inborn errors of metabolism by doing a metabolic screen test.

Q345

A patient has clinical features involved in 22q11.2 deletion syndrome but microarray is normal. Which gene might be involved?

ELN

PAFAH1B1

RAI1

TBX1

Q345 answer: TBX1

<https://pubmed.ncbi.nlm.nih.gov/11242110/>

<https://pubmed.ncbi.nlm.nih.gov/14585638/>

<https://pubmed.ncbi.nlm.nih.gov/16684884/>

<https://pubmed.ncbi.nlm.nih.gov/17273972/>

Q345 explanation

The TBX1 gene, a member of a conserved gene family that shares a common DNA-binding domain (the T-box), is located on chromosome 22q11.2. Pathogenic variants in this gene can result in at least some of the manifestations of 22q11.2 deletion syndrome. The other genes mentioned are involved in manifestations related to other contiguous gene/cytogenomic conditions (*ELN*: Williams syndrome; *PAFAH1B1* [*LIS1*]: Miller-Dieker syndrome; *RAI1*: Smith-Magenis syndrome).

Reference(s): Jerome and Papaioannou, Nat Genet 2001; Yagi et al., *Lancet* 2003; Paylor et al., *PNAS USA* 2006; Zweier et al., *Am J Hum Genet* 2007

ChatGPT first answer: TBX1

ChatGPT second answer: ELN

ChatGPT explanation:

The most likely gene involved in this scenario would be TBX1. This is because TBX1 is located within the 22q11.2 deletion region, and is considered a "critical gene" for the development of the clinical features associated with this syndrome. Mutations or deletions of TBX1 have been shown to be the most common genetic cause of this disorder. Microarray is a test that detect deletions or duplications in a genome, but not point mutations. Therefore, if the microarray is normal, this means there is no deletion or duplication of the 22q11.2 region, but there may still be a point mutation within a specific gene, like TBX1.

Q346

A patient undergoes genetic testing because of a strong family history of papillary renal cell carcinoma. Which of the following genes would most likely be indicated for inclusion in a genetic testing panel for this patient?

ANKRD26

MET

RET

STK11

Q346 answer: MET

<https://pubmed.ncbi.nlm.nih.gov/9731534/>

<https://pubmed.ncbi.nlm.nih.gov/15994935/>

<https://pubmed.ncbi.nlm.nih.gov/32561872/>

Q346 explanation

MET is a protooncogene; heterozygous pathogenic variants (“mutations”) in this gene have been described in hereditary papillary renal cell carcinoma, a type of hereditary renal cancer. Hereditary renal cancer may also occur related to variants in other genes, such as *FH*, *FLCN*, VHL, and others; cytogenomic abnormalities may also contribute to oncogenesis. In addition to hereditary papillary renal cell carcinoma, variants in the *MET* gene may also result in other genetic conditions.

Reference(s): Zhuang et al., Nat Genet 1998; Yang et al., Cancer Res 2005; Dizman et al., Nat Rev Nephrol 2020

Q347

Prenatal ultrasound shows findings consistent with Thanataphoric dysplasia; molecular testing via amniocentesis confirms the diagnosis. No parental testing has been done yet. What is the most likely recurrence risk?

<1%

25%

33%

50%

Q347 answer: <1%

<https://pubmed.ncbi.nlm.nih.gov/8843623/>

<https://www.ncbi.nlm.nih.gov/books/NBK1366/>

Q347 explanation

Thanataphoric dysplasia (TD) is a severe type of skeletal dysplasia, and is caused by heterozygous pathogenic variants (“mutations”) in the *FGFR3* gene. These variants usually occur de novo (though there is a theoretical possibility of issues such as germline mosaicism or other unusual situations that would affect the recurrence risk in a given situation). As the variants usually occur de novo, the risk to a subsequent conception would be about the same as in the general population, or far less than 1%. There are different types of TD, which are traditionally classified according to the specific skeletal findings.

Reference(s): Schild et al., Ultrasound Obstet Gynecol 1996; French and Savarirayan, GeneReviews 2020

ChatGPT first answer: 50%

ChatGPT second answer: <1%

ChatGPT explanation (the answer reverted back to 50%, an incorrect answer):

50%. Thanataphoric dysplasia (TD) is a rare genetic disorder that is caused by a mutation in the FGFR3 gene. The disorder is inherited in an autosomal dominant manner, meaning that an individual only needs to inherit one copy of the mutated gene to develop the disorder. The chance of passing the disorder to an offspring is 50% with each pregnancy.

When a prenatal ultrasound shows findings consistent with TD and molecular testing confirms the diagnosis, the recurrence risk for future pregnancies is 50%. It does not matter whether or not parental testing has been done yet, as the dominant inheritance pattern of TD means that if the fetus is affected, one of the parents must also be affected. It's worth noting that the recurrence risk may change if the specific genetic mutation that causes the TD is known and one of the parents is found to be a carrier of that specific mutation.

Q348

A patient is suspected to have mosaicism for a specific genetic variant (a rare single nucleotide variant). Which of the following assays would likely provide the most precise information about the level of mosaicism for this variant?

Digital droplet PCR

Genome sequencing

SNP microarray

NGS-based panel testing

Q348 answer: Digital droplet PCR

<https://pubmed.ncbi.nlm.nih.gov/10430926/>

<https://pubmed.ncbi.nlm.nih.gov/29655203/>

<https://pubmed.ncbi.nlm.nih.gov/29717444/>

<https://pubmed.ncbi.nlm.nih.gov/33108101/>

Q348 explanation

The clinical relevance of mosaic variants is becoming increasingly appreciated, including the fact that these variants can impact many different genes and clinical conditions. There are a number of different ways to assess mosaicism. Droplet digital polymerase chain reaction (ddPCR) enables the quantification of specifically targeted genetic variants via counting the proportion of the targeted nucleic acid molecules that are present in different “partitions” of the sample. ddPCR can be used to precisely detect a variety of types of genetic changes, including those present at levels too low for other methods to reliably detect. This method is used increasingly in research contexts and may also be available in some clinical scenarios.

Reference(s): Vogelstein et al., Proc Natl Acad Sci USA 1999; Lindy et al., Epilepsia 2018; Zhou et al., Methods Mol Biol 2018; Beck et al., N Engl J Med 2020

Q349

Genetics is consulted because a young patient has a hand malformation and ipsilateral pectoralis major hypoplasia. Which term refers to these findings?

Klippel-Feil anomaly

Pectus carinatum

Poland anomaly

Sprengel anomaly

Q349 answer: Poland anomaly

<https://pubmed.ncbi.nlm.nih.gov/20811224/>

<https://pubmed.ncbi.nlm.nih.gov/25633462/>

Q349 explanation

Poland anomaly refers to unilateral (one-sided) pectoralis major absence or hypoplasia with ipsilateral (same-sided) anomaly of the upper limb, especially affecting the hand. The condition is typically sporadic, but familial cases have been described. Poland anomaly can occur in an isolated fashion or can be observed in a syndromic context.

Reference(s): Human Malformations and Related Anomalies, 3rd ed.; Seyfer et al., *Plast Reconstr Surg* 2010; Yiyit et al., *Ann Thorac Surg* 2015

Q350

Due to family history and genetic testing in affected relatives, a healthy patient is suspected to have a balanced translocation. Which test would be most likely to help efficiently identify this translocation?

Karyotype

NGS-based panel testing

Sanger sequencing

SNP-based microarray

Q350 answer: Karyotype

<https://pubmed.ncbi.nlm.nih.gov/29956287/>

Q350 explanation

Though increasingly replaced by newer molecular technologies in a variety of clinical contexts, the karyotype can be an useful, efficient, and available assay to visualize the chromosomes in certain scenarios. The karyotype can be used to detect balanced translocations; some other technologies, such as the other choices mentioned will not typically be able to detect these translocations, either because they are too limited in scope in terms of the genetic/genomic information evaluated, or because they do not provide adequate information about the physical arrangement/location of genetic/genomic material.

Reference(s): Thompson & Thompson, Genetics in Medicine, 7th ed; Wilch and Morton, *Adv Exp Med Biol* 2018

Q351

A pathogenic variant in which of the following genes can cause a phenotype similar to deletion of chromosome 17p11.2?

NF1

RAI1

RBM8A

TBX1

Q351 answer: RAI1

<https://pubmed.ncbi.nlm.nih.gov/11404004/>

<https://pubmed.ncbi.nlm.nih.gov/12652298/>

<https://www.ncbi.nlm.nih.gov/books/NBK1310/>

Q351 explanation

Smith-Magenis syndrome (SMS), which involves intellectual disability, behavioral differences, and distinct craniofacial and skeletal findings, is caused by heterozygous deletion of chromosome 17p11.2. Heterozygous pathogenic variants in the gene *RAI1*, which is located in the SMS chromosomal region, can also cause the condition.

Reference(s): Seranski et al., Gene 2001; Slager et al., Nat Genet 2003; Smith et al., GeneReviews 2019

ChatGPT first answer: NF1

ChatGPT second answer: NF1

ChatGPT explanation (when asked the question and to provide an explanation, the correct answer was given):

RAI1. A pathogenic variant in the RAI1 gene can cause a phenotype similar to deletion of chromosome 17p11.2.

RAI1 (Retinoic acid-induced 1) gene is located on chromosome 17p11.2, and it has been reported that variations in this gene can lead to similar features as those seen in deletion of 17p11.2, which is a common genetic disorder. The deletion of this region results in loss of the RAI1 gene, leading to a wide range of symptoms, including developmental delay, intellectual disability, speech impairment, and behavioral problems.

A pathogenic variant in RAI1 can cause Smith-Magenis syndrome (SMS), which is characterized by developmental delay, intellectual disability, craniofacial anomalies, sleep disturbances, and behavioral problems. SMS is caused by a deletion of 17p11.2, which encompasses the RAI1 gene.

It's worth noting that deletion of 17p11.2 can also cause other genetic disorders such as Potocki-Shaffer syndrome, and other genes located on this region can contribute to the symptoms, but RAI1 is considered the most likely gene in this scenario.

Q352

A patient has a family history that is highly suspicious for Brugada syndrome. Assuming the clinical suspicion is correct, what gene is most likely to be involved?

CACNA1C

SCN1B

SCN5A

SEMA3A

Q352 answer: SCN5A

<https://www.ncbi.nlm.nih.gov/books/NBK1517/>

<https://pubmed.ncbi.nlm.nih.gov/29959160/>

<https://pubmed.ncbi.nlm.nih.gov/32893267/>

Q352 explanation

Multiple genes have been implicated in Brugada syndrome, a cardiac arrhythmia syndrome that involves risk of sudden death. Recent evaluation of variants has called certain genes and variants into question in terms of causation, but the *SCN5A* gene is the most frequently involved gene in individuals with Brugada syndrome. Many different genes are involved in various cardiac arrhythmia disorders.

Reference(s): Brugada et al., GeneReviews 2016; Hosseini et al., Circulation 2018; Walsh et al., Genet Med 2021

Q353

A man and a woman are seen together for prenatal counseling. The couple, both of whom are healthy, are from a community where a childhood-onset autosomal recessive genetic condition affects about 1 in 100 people. The man’s brother has the condition. What is the approximate chance that the couple’s child will be affected?

0.1%

1%

3%

6%

Q353 answer: 3%

Q353 explanation

Several calculations have to be made to arrive at the answer. There are some shortcuts/estimates that can be made to arrive at an approximate answer, but for the sake of education, we will not use these shortcuts in the explanation here. First, we know that the man has a brother who has the condition. The man is healthy, so he does not have the condition. There is therefore a ⅔ chance he is a carrier, and a ⅓ chance he is not a carrier (if we didn’t know whether he was affected, there would be a ½ (50%) chance he would be a carrier, but we know his clinical status, so we can eliminate the possibility that he is affected when doing the calculation). The condition affects 1/100 people in the population, so the frequency of the recessive allele (q) is 1/10, while the frequency of the wild-type allele (p) is 9/10. We estimate the prevalence of carriers as 2pq, or (2)(9/10)(1/10) = 18/100. However, just to be precise, since we know that the woman does not have the condition, we can say that the chance of her being a carrier is 18/99. This is the same kind of adjustment we made previously, though it doesn’t affect the final result much at all, and often won’t be necessary to arrive at a final estimate in this type of calculation. Now, we know that the chance of the man being a carrier is ⅔ and the chance of the woman being a carrier is 18/99. The chance of them both passing the affected alleles on to their child is ¼. Lastly, we multiply these 3 fractions together to get the final answer: (⅔)(18/99)(¼) = approximately 3%.

Reference(s): Thompson & Thompson, Genetics in Medicine, 8th ed

Q354

A patient with neonatal-onset epilepsy has been found to have bi-allelic pathogenic variants in the gene *ALDH7A1*. What treatment might be effective based on the genetic information?

Allopurinol

Colchicine

Levodopa

Pyridoxine

Q354 answer: Pyridoxine

<https://pubmed.ncbi.nlm.nih.gov/16491085/>

<https://pubmed.ncbi.nlm.nih.gov/25192505/>

<https://pubmed.ncbi.nlm.nih.gov/33200442/>

Q354 explanation

There are many different genetic causes of seizure disorders. Sometimes, knowing the molecular genetic cause can be especially important to determine the optimal treatment, which can in turn significantly impact morbidity and mortality. Individuals with bi-allelic pathogenic variants in *ALDH7A1* may have pyridoxine-responsive epilepsy. The condition manifests with neonatal-onset seizures, and is unresponsive to standard antiepileptic drugs, but, as the name implies, will respond to treatment with pyridoxine. Adjunctive therapy (lysine reduction therapies) has also been recommended for patients with this condition.

Reference(s): Mills et al., Nat Med 2006; Olson et al., Semin Neurol 2014; Coughlin et al., J Inherit Metab Dis 2021

Q355

A patient seen by genetics is noted to have developmental delay, obesity, postaxial polydactyly, and renal disease. What condition is most likely?

Bardet-Biedl syndrome

Joubert syndrome

McKusick-Kaufman syndrome

Senior-Loken syndrome

Q355 answer: Bardet-Biedl syndrome

<https://pubmed.ncbi.nlm.nih.gov/20301537/>

<https://pubmed.ncbi.nlm.nih.gov/26762677/>

Q355 explanation

Bardet-Biedl syndrome (BBS), which involves manifestations affecting multiple organ systems, is a type of ciliopathy. Characteristic features of BBS include developmental delay/intellectual disability, obesity, postaxial polydactyly, and renal disease, as well as other features, such as distinctive craniofacial features, and genitourinary anomalies. Pathogenic variants in multiple genes have been identified as causing BBS. There are also a number of other genetic conditions that are considered ciliopathies, and many of these can have overlapping features with BBS.

Reference(s): Khan et al., *Clin Genet* 2016; Forsyth & Gunay-Aygun, *GeneReviews* 2020; Jones, Smith’s Recognizable Patterns of Human Malformation, 7th ed.

Q356

An apparently healthy adult woman is referred to genetics because her father and sister have what appears to be a type of familial hypertrophic cardiomyopathy. Who would be the most informative family member to test first?

The woman’s child

The woman’s father

The woman herself

The woman’s mother

Q356 answer: The woman’s father

<https://www.ncbi.nlm.nih.gov/books/NBK1768/>

Q356 explanation

In this question, we know that the father and sister have familial hypertrophic cardiomyopathy, but the genetic cause has not yet been identified. The woman may have been referred because she has concerns about her health or that of other family members who may be affected (such as her children). In this situation, although the woman is described as healthy, we do not know for sure whether she harbors a pathogenic variant that may be causing the condition in her father and sister. Genetic testing of her father (or sister) might identify the genetic cause in the family, which could potentially allow genetic testing in other family members to be considered at that point. If one were to start by testing the woman herself (prior to testing family members who are clearly affected), interpreting the results might be more difficult. These situations arise frequently in clinical practice (as well as research-related situations).

Reference(s): Thompson & Thompson, Genetics in Medicine, 8th ed; Cirino and Ho, GeneReviews 2021

ChatGPT first answer: The woman herself

ChatGPT second answer: The woman herself

ChatGPT explanation:

The woman herself. Familial hypertrophic cardiomyopathy (FHC) is an inherited disorder of the heart muscle that is caused by mutations in various genes, including MYH7, MYBPC3, TNNI3, TNNT2, TPM1, ACTC1, and others. FHC is inherited in an autosomal dominant manner, which means that an individual only needs to inherit one copy of the mutated gene to develop the disorder.

In this scenario, the woman's father and sister have what appears to be a type of familial hypertrophic cardiomyopathy. The most informative family member to test first would be the woman herself, as it will determine whether she carries the genetic mutation that causes the disorder. If the woman is found to have the same genetic mutation as her father and sister, it will confirm the diagnosis of FHC in the family and will provide information about the recurrence risk for future pregnancies. If the woman does not carry the genetic mutation, it would suggest that the disorder may not be inherited in an autosomal dominant manner, or that the genetic mutation has not been identified yet.

It's worth noting that testing the woman's mother could also be informative, as she may be a carrier of the genetic mutation, and this could help identify the specific genetic mutation that is causing the disorder in the family.

Q357

An apparently healthy adult woman was referred to genetics because her father and sister have familial hypertrophic cardiomyopathy. Genetic testing reveals a VUS in a relevant gene in the father, which is not resolved by further familial testing. Based on this, what is recommended for the woman?

Discharge from care

Genetic testing of the woman’s children

Immediate ICD placement

Regular clinical cardiac evaluation

Q357 answer: Regular clinical cardiac evaluation

<https://www.ncbi.nlm.nih.gov/books/NBK1768/>

Q357 explanation

In the description of the family, a variant of uncertain significance (VUS) has been identified, but its pathogenicity has not been resolved by further familial testing. Based on this information, the woman would be advised to continue to undergo regular clinical cardiac evaluation; this evaluation plan would likely be tailored to the specific genetic condition and family history. Clinical findings (e.g., if cardiac testing were to identify signs of cardiomyopathy) would be treated accordingly. This advice might change if new data emerged that enabled the variant to be reclassified.

Reference(s): Thompson & Thompson, Genetics in Medicine, 8th ed; Cirino and Ho, *GeneReviews* 2021

Q358

A several month-old boy has a scaly skin rash. Assuming a genetic diagnosis, deficiency of what enzyme might be suspected?

Aldolase B

Arginase

Mevalonate kinase

Steroid sulfatase

Q358 answer: Steroid sulfatase

<https://pubmed.ncbi.nlm.nih.gov/10583107/>

[Genetics of Inherited Ichthyoses and Related Diseases](https://pubmed.ncbi.nlm.nih.gov/32147747/)

Q358 explanation

Steroid sulfatase deficiency, which results from pathogenic variants affecting the *STS* gene, is an X-linked condition sometimes referred to as X-linked ichthyosis. The condition can also include corneal opacities, cryptorchidism, and neurological manifestations. There are many other genetic conditions that also cause ichthyosis.

Reference(s): Martin et al., Br J Dermatol 1999; Fischer and Bourrat, Acta Derm Venereol 2020

Q359

Which of the following refers to the most common isochromosome in humans?

i(1q)

i(18p)

i(Xq)

i(Yp)

Q359 answer: i(Xq)

Q359 explanation

The term “isochromosome” refers to a chromosome that results from duplication of part of a chromosome and deletion of the non-duplicated part. This yields a chromosome that is a mirror image copy of the duplicated chromosomal region.  There are multiple types of isochromosomes; the type depends on where on the chromosome the chromosomal break occurs. The most common isochromosome affects the long arm of the X chromosome, and is written using standard nomenclature as i(Xq).

Reference(s): Gonzaga-Jauregui and Lupski, Genomics of Rare Diseases

ChatGPT first answer: i(18p)

ChatGPT second answer: i(18p)

ChatGPT explanation (when asked the question and to provide an explanation, the correct answer was given):

i(Xq). An isochromosome is a type of structural chromosome abnormality that is formed when one arm of a chromosome is lost and the other arm is duplicated, resulting in two identical arms of the same length. The most common isochromosome in humans is i(Xq), which is formed when the short arm (p) of the X chromosome is lost and the long arm (q) is duplicated.

Isochromosome Xq is the most common isochromosome in humans because it is formed during meiosis when the X chromosome is not paired with a Y chromosome. This results in the X chromosome being pulled to the same pole of the cell and the formation of an isochromosome. This isochromosome can be seen in Turner syndrome, a chromosomal disorder that affects females, characterized by short stature, webbed neck, and gonadal dysgenesis (absence or malformation of ovaries).

It's worth noting that each of the other options represents a different type of isochromosome: i(1q) is a rare isochromosome that can be associated with malformations, i(18p) is a rare isochromosome that can be associated with developmental delay, and i(Yp) is a rare isochromosome that can be associated with Turner Syndrome in males.

Q361

The Genetic Information Nondiscrimination Act (GINA) of 2008 aims to protect Americans from discrimination based on their genetic information related to which of the choices below?

Disability insurance and Life insurance

Employment

Health insurance and Employment

Long-term care insurance

Q361 answer: Health insurance and employment

<https://www.genome.gov/about-genomics/policy-issues/Genetic-Discrimination>

<https://pubmed.ncbi.nlm.nih.gov/19348693/>

<https://pubmed.ncbi.nlm.nih.gov/34400812/>

Q361 explanation

A number of different laws help guide and regulate issues related to genetics and genomics, including how this information can be used in different situations. In the United States, the Genetic Information Nondiscrimination Act (GINA), enacted in 2008, helps provide protection from discrimination based on a person’s genetic information as relates to health insurance and employment.

Reference(s): NHGRI, Policy Issues in Genomics, Genetic Discrimination; McGuire and Majumder, Genome Med 2009; Lenartz et al., Genet Med 2021

Q363

A woman is affected by an autosomal dominant condition (that is completely penetrant). She has three children. What is the chance that all three of her children are affected by the condition?

12.5%

25%

50%

75%

Q363 answer: 12.5%

Q363 explanation

The woman has an autosomal dominant condition; each of her children has a 50% (0.5) chance of inheriting the causative allele. Since she has 3 children, the chance of all of them being affected can be calculated as (0.5)(0.5)(0.5) = 0.125, or 12.5%.

Reference(s): Thompson & Thompson, Genetics in Medicine, 8th ed.

ChatGPT first answer: 25%

ChatGPT second answer: 25%

ChatGPT explanation:

25%. An autosomal dominant condition is one in which an individual only needs to inherit one copy of the mutated gene to develop the disorder. If the woman is affected by an autosomal dominant condition, she has a 50% chance of passing the genetic mutation to each of her children.

The chance that all three of her children are affected by the condition is (1/2)^3 = 1/8 = 12.5%.

It's worth noting that this probability assumes that the condition is completely penetrant, meaning that all individuals who carry the genetic mutation will develop the disorder, and also assumes that there is no genetic variation such as incomplete penetrance or variable expressivity, in which case the chance that all three children are affected could vary. Also, assuming that the woman's partner is not a carrier of the genetic mutation and therefore the chance of all children being affected is 12.5%

Q364

Due to physical examination and family history findings suggestive of a genetic condition, a person has been found to have a pathogenic variant in *FBN1*. Based on this, what medical treatment would most likely be indicated (though this would only be indicated for some conditions related to this gene)?

Anakinra

Beta blocker

Carnitine

Digoxin

Q364 answer: Beta blocker

<https://www.ncbi.nlm.nih.gov/books/NBK1335/>

<https://pubmed.ncbi.nlm.nih.gov/34475413/>

Q364 explanation

Marfan syndrome, as well as multiple other conditions, can be caused by pathogenic variants in the FBN1 gene. Marfan syndrome affects multiple organ systems, including the eyes, musculoskeletal system, and cardiovascular system. Among other management considerations, medical management of cardiovascular sequelae with beta blockers and/or angiotensin receptor blockers is often used in order to help reduce hemodynamic stress on the aortic wall.

Reference(s): Dietz, GeneReviews 2017; Milewicz et al., Nat Rev Dis Primers 2021

Q365

Based on medical history and examination, a patient is suspected to have Pallister-Killian syndrome. What tissue source(s) or sample type(s) would likely be most helpful to confirm the diagnosis?

CSF

Peripheral blood or saliva

Buccal swab or skin biopsy

Urine

Q365 answer: Buccal swab or skin biopsy

<https://pubmed.ncbi.nlm.nih.gov/10955474/>

<https://pubmed.ncbi.nlm.nih.gov/30140312/>

<https://pubmed.ncbi.nlm.nih.gov/30430791/>

Q365 explanation

Pallister-Killian syndrome, sometimes called Pallister-Killian mosaic syndrome, is caused by mosaic tetrasomy of chromosome 12p (typically involving an additional isochromosome 12p). Manifestations are variable, but can include neurocognitive impairment, seizures, and distinctive features on physical examination, as well as other findings. Blood-based chromosomal testing will typically not detect the causative cytogenomic anomalies, but testing via buccal swab (which is easier to conduct, but involves some limitations) or skin biopsy can identify the cytogenomic cause.

Reference(s): Manasse et al., Clin Dysmorphol 2000; Karaman et al., Mol Cytogenet 2018; Bertini et al., Ann Lab Med 2019

ChatGPT first answer: Peripheral blood or saliva

ChatGPT second answer: Buccal swab or skin biopsy

ChatGPT explanation (ChatGPT explanation (the answer reverted back to Peripheral blood or saliva%, an incorrect answer):

Peripheral blood or saliva. Pallister-Killian syndrome (PKS) is a rare genetic disorder caused by an extra genetic material on the short arm of chromosome 12, specifically at the 12p11.2-12p13.1 locus. The extra genetic material is usually caused by an isochromosome, which is a type of structural chromosome abnormality. The most common method of diagnosis is by karyotyping, which is a cytogenetic method that looks at the number and structure of chromosomes in a cell. The most informative tissue source to obtain the chromosomes would be peripheral blood or saliva.

A peripheral blood sample, is a common and non-invasive source of chromosomes, which can be analyzed by karyotyping, which is a cytogenetic method that looks at the number and structure of chromosomes in a cell. A saliva sample can also be used for DNA extraction, and then performing chromosomal microarray analysis (CMA) to detect chromosomal abnormalities.

It's worth noting that other tissue sources such as skin biopsy, urine, and CSF sample would not be informative for diagnosis of PKS because they do not contain chromosomes. Buccal swab sample could be used to extract DNA and confirm the diagnosis but it is not the most informative tissue source.

Q366

For which condition is there an FDA-approved gene therapy available?

ADA-SCID

Familial hypercholesterolemia

Progeria

Spinal muscular atrophy

Q366 answer: Spinal muscular atrophy

<https://pubmed.ncbi.nlm.nih.gov/29326244/>

<https://pubmed.ncbi.nlm.nih.gov/31365802/>

Q366 explanation

Gene therapies have been developed and tested to treat a number of genetic conditions, including a type of spinal muscular atrophy (SMA), which was FDA-approved in 2019. Gene therapy and gene editing-based treatments are being studied related to many other conditions. Of note, gene therapy for ADA-SCID has been approved in Europe, but not by the FDA in the US.

Reference(s): Dunbar et al., Science 2018; High and Roncarlo, New Engl J Med 2019

Q367

Which of the following refers to a sequence of DNA that closely resembles a gene, but which has become (apparently) inactive through evolution, and which can result in challenges in genetic testing?

Euchromatin

Haplotype

Pseudogene

Recombinant

Q367 answer: Pseudogene

<https://www.genome.gov/genetics-glossary/Pseudogene>

Q367 explanation

A pseudogene is a sequence of DNA that closely resembles a gene, but which has become (apparently) inactive through evolution. The presence of pseudogenes can cause challenges in genetic testing processes; for example, it can be difficult to determine if a detected variant occurs in a gene or a related pseudogene.

Reference(s):<https://www.genome.gov/genetics-glossary/Pseudogene>

Q369

A child with optic atrophy has been diagnosed with Wolfram syndrome. Testing for what other clinical manifestation would be warranted?

Diabetes mellitus

Hirschsprung disease

Hypercholesterolemia

Immunodeficiency

Q369 answer: Diabetes mellitus

<https://pubmed.ncbi.nlm.nih.gov/20301750/>

<https://pubmed.ncbi.nlm.nih.gov/33529164/>

Q369 explanation

Wolfram syndrome is an autosomal recessive condition caused by pathogenic variants in *WFS1*. The condition can involve multiple manifestations, including optic atrophy, hearing loss, diabetes mellitus and other endocrine abnormalities, and neurodegeneration, as well as other findings.

Reference(s): Tranebjærg et al., GeneReviews 2020; Zhang et al., J Clin Invest 2021

Q370

A neonate has an older brother with molecularly-confirmed familial hemophagocytic lymphohistiocytosis (fHLH). Based on the causes of fHLH, what is the most likely chance that the neonate will also be “genetically affected” (have also inherited the causative variant(s))?

25%

33%

50%

67%

Q370 answer: 25%

<https://pubmed.ncbi.nlm.nih.gov/20301617/>

Q370 explanation

Familial hemophagocytic lymphohistiocytosis (fHLH) is an autosomal recessive immunologic disorder that can be caused by pathogenic variants in multiple known genes. (However, there are rare reports of affected individuals found to have heterozygous variants of one gene). Excluding these rare reports, the history of an older affected sibling means that this infant would be expected to have a 25% chance of also being affected. fHLH typically presents with fever, hepatosplenomegaly, and cytopenia. Accurate diagnosis is important for management.

Reference(s): Zhang et al., GeneReviews 2021

ChatGPT first answer: 50%

ChatGPT second answer: 50%

ChatGPT explanation (explanation was truncated/ended by ChatGPT prior to completion):

50%. Familial Hemophagocytic Lymphohistiocytosis (fHLH) is a rare genetic disorder caused by mutations in various genes involved in the immune system, including PRF1, UNC13D, STX11, STXBP2, RAB27A and LYST. The disorder is inherited in an autosomal recessive manner, meaning that an individual must inherit two copies of the mutated gene, one from each parent, in order to develop the disorder.

In this scenario, the neonate has an older brother with molecularly-confirmed fHLH, which means that the parents of the neonate are likely carriers of the genetic mutation that causes the disorder. The chance that the neonate will also be "genetically affected" (have also inherited the causative variant(s)) is 50%. Each parent has a 50% chance of passing the genetic mutation to the neonate, and since both parents are carriers, the chance that the neonate will inherit the genetic mutation from both parents is (1/2) x (1/2) = 1/4 = 25%.

It's worth noting that these probabilities are based on the assumption that the parents are carriers of the same genetic mutations that cause the disorder and assuming Mendelian inheritance pattern. If the parents are carriers of different mutations in the same

Q371

Overall in humans, autosomal aneuploidies most commonly occur due to events during which stage of meiosis?

Maternal meiosis I

Maternal meiosis II

Paternal meiosis I

Paternal meiosis II

Q371 answer: Maternal meiosis I

<https://pubmed.ncbi.nlm.nih.gov/11283700/>

Q371 explanation

Human autosomal aneuploidies are relatively common. Overall, the meiotic event giving rise to these aneuploidies most commonly occurs during maternal meiosis I. However, the proportion varies according to the particular aneuploidy; depending on the aneuploidy, events may also frequently occur during maternal meiosis II or during paternal meiosis I or II or during postzygotic mitosis.

Reference(s): Hassold and Hunt, Nat Rev Genet 2001

ChatGPT first answer: Maternal meiosis II

ChatGPT second answer: Maternal meiosis I

ChatGPT explanation:

Maternal meiosis I. Overall in humans, autosomal aneuploidies most commonly occur due to events during maternal meiosis I. During meiosis, homologous chromosomes pair up and exchange genetic material through a process called crossing over. This can lead to abnormal distribution of chromosomes, resulting in aneuploidy. Nondisjunction, which is when chromosomes fail to separate properly, is the most common type of aneuploidy.

During maternal meiosis I, errors in chromosomal separation are more common due to the age of the mother. As a woman ages, the risk of errors in meiosis increases, leading to a higher risk of aneuploidies in her eggs. This is why, overall, autosomal aneuploidies most commonly occur due to events during maternal meiosis I.

It's worth noting that there are also other factors that can increase the risk of aneuploidy such as exposure to certain environmental toxins, and genetic disorders such as Turner syndrome, Down syndrome, and Klinefelter syndrome.

Q373

You are seeing a patient suspected to have septo-optic dysplasia. In addition to agenesis/anomalies of the septum pellucidum and corpus callosum and optic nerve hypoplasia, what finding would you most likely anticipate?

Hirschsprung disease

Melanoma

Pituitary anomalies

Rhizomelia

Q373 answer: Pituitary anomalies

<https://pubmed.ncbi.nlm.nih.gov/21037540/>

<https://pubmed.ncbi.nlm.nih.gov/34238479/>

Q373 explanation

Septo-optic dysplasia (SOD) includes the triad of agenesis/anomalies of the septum pellucidum and corpus callosum, optic nerve hypoplasia, and pituitary/hypothalamic anomalies. Other clinical features may be present as well. Pathogenic variants in multiple genes have been reported as causative of SOD.

Reference(s): Fard et al., Pediatr Endocrinol Rev 2010; Sataite et al., Handb Clin Neurol 2021

Q374

A woman and a man have a child. Later, the man has another child, this time with the woman’s sister. Approximately what proportion of their DNA would the two children be expected to have in common?

25%

37.5%

50%

67%

Q374 answer: 37.5%

<https://pubmed.ncbi.nlm.nih.gov/33452467/>

Q374 explanation

On average, full siblings have about 50% of their DNA in common. Half-siblings have about 25% of their DNA in common. In this question, the children have the same father and their mothers are sisters; these children have been termed “three-quarter siblings.” These children would be expected, on average, to share about 37.5% of their DNA.

Reference(s): Galván-Femenía et al., Heredity (Edinb) 2021

ChatGPT first answer: 25%

ChatGPT second answer: 50%

ChatGPT explanation:

50%. The proportion of DNA that two individuals share is determined by their degree of relatedness. The two children are half-siblings, which means they share one biological parent (the man) and therefore they would be expected to have 50% of their DNA in common.

Half-siblings share, on average, 25% of their DNA, as they share only one parent. The proportion of DNA that they share can vary slightly due to the randomness of which segments of DNA are inherited from each parent, but on average they share 25%.

It's worth noting that these probabilities are based on Mendelian inheritance pattern, and assuming no consanguinity, and no genetic variation such as mutation. Also, the percentage of shared DNA is an estimate and the actual amount of shared DNA can vary based on the specific segments inherited from each parent.

Q375

You are asked to do a genetics consult for a patient whose condition includes overgrowth, vascular malformations, and skin findings (eg, epidermal nevus). Genome sequencing on peripheral blood was unrevealing. Which gene might you suspect?

GNAQ

PIK3CA

PTPN11

TP53

Q375 answer: PIK3CA

<https://pubmed.ncbi.nlm.nih.gov/25557259/>

<https://pubmed.ncbi.nlm.nih.gov/34854542/>

Q375 explanation

*PIK3CA*-Related Overgrowth Spectrum (PROS) includes a variety of overlapping phenotypes that usually result from postzygotic/somatic mutation of the *PIK3CA* gene. These phenotypes have previously been given different terms; frequent features of PROS phenotypes can include:  hemihyperplasia, overgrowth, vascular anomalies, skin abnormalities, neoplasms, scoliosis, and other findings.

Reference(s): Keppler-Noreuil et al., Am J Med Genet A 2015; Filippides et al., Am J Med Genet A 2021

Q377

An adult woman has a history of easy bruising and prolonged bleeding. Her father had acute myelogenous leukemia; she mentions that he also seemed to have similar bruising and bleeding issues. What gene would most likely be involved?

PTEN

PTCH1

RUNX1

VHL

Q377 answer: RUNX1

<https://pubmed.ncbi.nlm.nih.gov/32315381/>

<https://www.ncbi.nlm.nih.gov/books/NBK568319/>

Q377 explanation

Heterozygous germline pathogenic variants in the gene *RUNX1* can result in a condition termed *RUNX1* familial platelet disorder with associated myeloid malignancies (*RUNX1*-FPDMM). The condition can involve easy or abnormal bruising and prolonged bleeding (due to thrombocytopenia and platelet dysfunction), as well as increased risk of hematologic malignancy.

Reference(s): Simon et al., Blood 2020; Deuitch et al., GeneReviews 2021

Q379

A genetic condition that affects about 1 in 50,000 infants is added to newborn screening. The assay for the condition has 99.9% sensitivity and 99.9% specificity. An infant has a positive screen result. Based on just this information, what is the (approximate) chance the child has the condition?

0.01%

2%

98%

99.9%

Q379 answer: 2%

<https://pubmed.ncbi.nlm.nih.gov/10891024/>

<https://pubmed.ncbi.nlm.nih.gov/28613617/>

Q379 explanation

This question is asking for the positive predictive value (PPV) of the test. In this case, PPV refers to the chance that an infant with a positive test has the condition that has been added to newborn screening. The PPV depends on the prevalence of the condition in the tested population, as well as characteristics of the assay (sensitivity and specificity). The PPV of the test can be calculated by dividing the number of true positives (TP) by the total number of positives from the testing. The total number of positives includes both true positives as well as false positives. In other words, this can be expressed at: PPV = TP/(TP+FP). For a quick shortcut, PPV can be calculated using the following formula: PPV = (sensitivity x prevalence) / [ (sensitivity x prevalence) + ((1 – specificity) x (1 – prevalence))]. There are several online calculators that can be used to calculate this. Based on the information in the question, the chance that the infant really has the condition is about 2%. Another way to calculate the PPV is by looking at a possible scenario. Let’s say about 1 million infants are born each year. Since the condition affects about 1 in 50,000 infants, we would expect about 20 infants with the condition to be born each year. We know that the test is 99.9% sensitive - this means that 99.9% of people with the condition will have a positive test. So, we would estimate that 0.999*20, or 19.98 people with the condition would have a positive test. For our question, we can just round this to 20, though please note that it can sometimes be important not to use this rounding shortcut. Now, we also know that the test is 99.9% specific, meaning 99.9% of people without the condition will have a negative result, while 0.1% will have a false positive result. So, just to be precise, we can say that, of the 1 million infants in this population, all but 20 infants don’t have the condition: 1,000,000 - 20 = 999,980 (again, this step isn’t necessary for us to estimate the answer in this particular case, but let’s include it, just to be clear). The number of false positives in this population can be calculated by figuring out what is 0.1% of this number: 0.001*999,980= 999.98. We can again round this to 1,000 (but remember, there are times when it’s better not to round). So, we have about 20 true positives, and about 1,000 false positives. This means that the PPV is about 20 (true positives) out of about 1,020 (all positives), which is about 2%. As this example shows, for rare conditions, false positive results may be frequent (i.e., a positive screen result may be found much more often in unaffected people than in affected people). Because of this, follow-up testing after a positive screening result is often recommended. These concepts are important to understand when considering what the results of a test may mean.

Reference(s): Kwon and Farrell, Arch Pediatr Adol Med 2000; Tenny and Hoffman, StatPearls 2021

ChatGPT first answer: 98%

ChatGPT second answer: 2%

ChatGPT explanation:

2%. The chance that an infant with a positive screen result for a genetic condition actually has the condition is known as the positive predictive value (PPV). The PPV depends on the prevalence of the condition in the population, the sensitivity and specificity of the screening test, and the probability of a false positive result.

The formula for PPV is: PPV = (True positives) / (True positives + False positives)

Given that the assay for the condition has 99.9% sensitivity and 99.9% specificity, and the condition affects about 1 in 50,000 infants, we can calculate the PPV as follows: PPV = (1 in 50,000 x 0.999) / (1 in 50,000 x 0.999 + (1-1 in 50,000) x (1-0.999)) = 2%

It's worth noting that this is an approximate chance and it's based on the assumption that the population screened is representative of the population at risk and that the test performance is consistent with the given values. Also, a positive screen result doesn't necessarily mean the child has the condition, it needs to be confirmed by additional testing.

Q380

A man and a woman both have a clinical condition because they both have a certain heterozygous genetic variant. They have a child, who inherits both genetic changes (one from each parent). The child is much more severely affected than the parents. What is the term for this inheritance?

Codominance

Incomplete dominance

Variable expressivity

X-linked inheritance

Q380 answer: Incomplete dominance

<https://www.ncbi.nlm.nih.gov/books/NBK174884/>

<https://www.ncbi.nlm.nih.gov/books/NBK1152/>

Q380 explanation

Incomplete dominance (also sometimes called semidominance) refers to a situation where the heterozygote state is an intermediate between the two homozygote (or compound heterozygote, for an affected individual) states. For example, related to the question, a person without any of the mentioned genetic changes would be unaffected - they would have two “wild-type” alleles. A person with one genetic change (a heterozygote) might be mildly or moderately affected. A person with two genetic changes (a homozygote, who has bi-iallelic genetic changes) would be more severely affected. Although this may seem like an unusual situation, there are many examples of this in clinical genetics. For example, people with achondroplasia (who have heterozygous genetic changes in the gene *FGFR3*) are less affected than a person who is homozygous for these genetic changes in *FGFR3*. As another example, having bi-allelic pathogenic variants in genes that involve cholesterol metabolism (like *APOB*, *LDLR*, or *PCSK9*) results in more severe manifestations than having a heterozygous variant.

Reference(s): Thompson and Thompson, Genetics in Medicine, 8th ed; Youngblom et al., GeneReviews 2016; Legare et al., GeneReviews 2020

ChatGPT first answer: Variable expressivity

ChatGPT second answer: Variable expressivity

ChatGPT explanation (when asked the question and to provide an explanation, the correct answer was given):

Incomplete dominance. Incomplete dominance is a term used to describe a situation where the phenotype of an individual who inherits two copies of a genetic variant (homozygosity) is a blend of the two variants rather than the complete expression of either one. The parents in this scenario both have the same genetic variant but only one copy each, and they have a milder form of the condition, but when the child inherits one copy from each parent, they have two copies of the genetic variant and have a more severe form of the condition.

It's worth noting that this case of inheritance is rare and most genetic conditions are inherited in a dominant or recessive manner, and it's not always easy to predict the effect of genetic variants on the phenotype. Also, the term "variable expressivity" refers to the phenomenon where different individuals with the same genetic variant can present with varying degrees of severity of the same condition, and X-linked inheritance refers to the inheritance pattern of a genetic variant located on the X chromosome.

Q382

A person has a very rare genetic condition that affects about 1 in 500,000 people. A test to detect this condition has 95% sensitivity and 99% specificity. What is the chance that the test will be negative for this individual?

1%

5%

95%

99%

Q382 answer: 5%

<https://pubmed.ncbi.nlm.nih.gov/29209603/>

Q382 explanation

The sensitivity of a test refers to the chance that the test will be positive in a person with the condition. For the situation in the question, this means that the test has a 95% chance of being positive for this person (who has the condition in question). This also means that there is a 5% chance that the result will be negative for this person (this would be termed a “false negative” result). Conversely, specificity refers to the chance that the test will be negative in a person without the condition. The prevalence of the condition is not used to calculate the answer (unlike calculations involving positive or negative predictive value).

Reference(s): Trevethan, Front Public Health 2017

ChatGPT first answer: 1%

ChatGPT second answer: 5%

ChatGPT explanation:

1%. The chance that a test will be negative for an individual is known as the negative predictive value (NPV). The NPV depends on the prevalence of the condition in the population, the sensitivity and specificity of the screening test, and the probability of a false negative result.

The formula for NPV is: NPV = (True negatives) / (True negatives + False negatives)

Given that the test has 95% sensitivity and 99% specificity, and the condition affects about 1 in 500,000 people, we can calculate the NPV as follows: NPV = (499,999 in 500,000 x 0.99) / (499,999 in 500,000 x 0.99 + (1 in 500,000) x (1-0.95)) = 1%

It's worth noting that this is an approximate chance, based on the assumption that the population screened is representative of the population at risk and that the test performance is consistent with the given values. Also, a negative test result doesn't necessarily mean the individual does not have the condition, it needs to be confirmed by additional testing.

Q383

A person undergoes enzymatic testing for beta-hexosaminidase A (HEX A, related to Tay-Sachs disease). The result shows absent enzyme activity. However, the person is not affected, and does not carry a disease-associated allele. What is the best explanation for this?

Allele dropout

Gene duplication

Nondisjunction

Pseudodeficiency

Q383 answer: Pseudodeficiency

<https://www.ncbi.nlm.nih.gov/books/NBK1218/>

Disorders related to the *HEXA* gene include Beta-Hexosaminidase A deficiency, GM2 Gangliosidosis, Type I, and Tay-Sachs Disease. The conditions can be diagnosed by enzymatic testing showing abnormally low HEX A activity and biallelic pathogenic variants in the *HEXA* gene. Enzymatic testing of affected individuals will show absent to near-absent beta-hexosaminidase A enzymatic activity (and normal or elevated beta-hexosaminidase B enzyme activity). Enzymatic activity can seem low due to the presence of specific genetic changes (which are more common in certain ancestral populations such as French Canadians or Ashkenazi), since these variants cause the enzyme to be unable to process the synthetic  substrates used in testing.This results in false positive enzyme testing results. Due to these factors, it has been recommended to conduct testing for pseudodeficiency alleles in certain populations and in certain situations.

Reference(s): Toro et al., GeneReviews 2020

Q384

Some genetic conditions have considerable genetic heterogeneity (the condition can be caused by variants in different genes). Which of the following conditions, all of which can involve predisposition to certain types of cancer, involves the most genetic heterogeneity?

Fanconi anemia

Li-Fraumeni syndrome

Tuberous sclerosis

Von Hippel-Lindau syndrome

Q384 answer: Fanconi anemia

<https://www.ncbi.nlm.nih.gov/books/NBK1401/>

Q384 explanation

Fanconi anemia is a multi-system condition that demonstrates considerable genetic heterogeneity. The condition, which can involve findings such as congenital anomalies, hematologic anomalies, and an increased risk of certain types of cancer, can be caused by pathogenic variants in over a dozen currently identified genes. Fanconi anemia can be diagnosed through chromosome breakage studies and/or molecular testing.

Reference(s): Mehta and Ebens, GeneReviews 2021

ChatGPT first answer: Li-Fraumeni syndrome

ChatGPT second answer: Li-Fraumeni syndrome

ChatGPT explanation:

Li-Fraumeni syndrome. Li-Fraumeni syndrome (LFS) is a rare genetic disorder that predisposes individuals to a wide range of cancers, including sarcomas, brain tumors, breast cancer, adrenocortical carcinomas and leukemia. LFS is caused by mutations in the TP53 tumor suppressor gene, and it has been estimated that the majority of LFS cases (~90%) are caused by mutations in the TP53 gene. However, recent studies have shown that other genes also contribute to LFS, such as CHEK2, ATM and ATRX, which means that LFS is a genetically heterogeneous condition.

Fanconi anemia, Tuberous sclerosis, and Von Hippel-Lindau syndrome are also genetic conditions that predispose individuals to certain types of cancer, but they are caused by mutations in specific genes and do not involve as much genetic heterogeneity as LFS. Fanconi anemia is caused by mutations in specific genes that are involved in DNA repair. Tuberous sclerosis is caused by mutations in two specific genes TSC1 and TSC2 which are involved in cell growth regulation. Von Hippel-Lindau syndrome is caused by mutations in the VHL gene which is involved in oxygen sensing and regulation of angiogenesis.

Q385

You order a microarray for a child with a neurobehavorial phenotype involving autism and developmental delay; he has no other apparent pertinent clinical findings. Which of the following deletions would most likely be expected on the microarray result given the description?

16p11.2

17q11.2

7q11.23

8q23.3-q24.11

Q385 answer: 16p11.2

<https://pubmed.ncbi.nlm.nih.gov/20466091/>

<https://pubmed.ncbi.nlm.nih.gov/31182824/>

<https://pubmed.ncbi.nlm.nih.gov/20301775/>

<https://pubmed.ncbi.nlm.nih.gov/33667823/>

Q385 explanation

Microarray-based testing has been considered a first-line test for children with intellectual disability, though microarray testing in many clinical circumstances  is now often being supplanted by approaches like exome or genome testing. Deletion of a region of chromosome 16p11.2 results in a variable condition that can include speech and language effects with autistic and other neurobehavioral features, as well as neuromotor effects. Other clinical features may also be present in some individuals. This deletion is considered one of the most common known genetic contributions to autism spectrum disorders (ASD).

Reference(s): Miller et al., Am J Hum Genet 2010; Srivastava et al., Genet Med 2019; Chung et al., Curr Opin Genet Dev 2021; Taylor et al., GeneReviews 2021

Q387

A woman has been diagnosed with breast cancer. Which of the following findings would most suggest that this could be related to a pathogenic variant in *BRCA1* or *BRCA2*?

No affected relatives

Onset at 80 years of age

Triple-negative status

Unilateral cancer

Q387 answer: Triple-negative status

<https://pubmed.ncbi.nlm.nih.gov/22144499/>

<https://www.ncbi.nlm.nih.gov/books/NBK1247/>

Q387 explanation

“Triple-negative” breast cancer refers to breast cancer without estrogen receptor (ER) and progesterone receptor (PR) expression and without human epidermal growth factor receptor 2 (HER2) overexpression. Triple-negative breast cancer tends to be more aggressive, and is relatively more common in people with breast cancer related to germline pathogenic variants in BRCA1 or BRCA2.

Reference(s): Mavaddat et al., Cancer Epidemiol Biomarkers Prev 2012; Petrucelli et al., GeneReviews 2022

ChatGPT first answer: Unilateral cancer

ChatGPT second answer: Unilateral cancer

ChatGPT explanation:

Unilateral cancer would most suggest that this could be related to a pathogenic variant in BRCA1 or BRCA2. BRCA1 and BRCA2 are genes that are known to increase the risk of breast and ovarian cancer. Women who carry a pathogenic variant in these genes have a higher risk of developing breast cancer, and the cancer is often unilateral (affecting only one breast). This is in contrast to a more typical breast cancer, which usually affects both breasts.

Q388

A person undergoes exome sequencing. Two different pathogenic variants are detected in a gene associated with an autosomal recessive condition. Which of the following would likely best determine the variants’ inheritance (to see if the variants are in cis or trans)?

Genome sequencing

Karyotype

Microarray testing

Parental testing

Q388 answer: Parental testing

<https://pubmed.ncbi.nlm.nih.gov/27657676/>

Q388 explanation

When people undergo genetic testing, two different variants in the same gene may be identified. It can be unclear whether the two variants are in cis (are on the same copy of the gene on one chromosome) or are in trans (are each on a different copy of the gene, with one variant on each chromosome). One way to help determine this is to conduct parental testing. For example, if one variant is identified in the person’s mother, and one variant is identified in the person’s father, the variants would be said to be in trans, and have been inherited in such a way that - assuming they are both pathogenic variants - the person could be affected by an autosomal recessive condition related to the gene. On the other hand, if one parent also has both variants, the variants would be said to be in cis, and the person would typically be a “carrier” of the variants. There are exceptions to this explanation, and there can be other ways (such as lab-based methods using the proband sample) besides parental testing to determine the configuration of variants.

Reference(s): Evans and Jarvik,Genet Med 2017; Thompson & Thompson, Genetics in Medicine, 8th ed

Q390

Based on clinical history and previous testing, you suspect that a patient has ornithine transcarbamylase (OTC) deficiency. What biochemical test might help confirm this diagnosis?

Blood succinylacetone

CSF guanidinoacetate

Total carnitine

Urine orotic acid

Q390 answer: Urine orotic acid

<https://pubmed.ncbi.nlm.nih.gov/14705115/>

<https://www.ncbi.nlm.nih.gov/books/NBK154378/>

Q390 explanation

Ornithine transcarbamylase (OTC) deficiency is an X-linked inborn error of metabolism (IEM) caused by pathogenic variants in *OTC*. The condition may be suspected due to patient medical history, as well as suggestive laboratory findings, such as hyperammonemia, as well as high glutamine and low-to-normal citrulline on biochemical testing. The condition can be diagnosed by identifying a causative genetic change in *OTC*, or by increased orotic acid excretion in a random urine collection or after an allopurinol challenge. In males, decreased hepatic OTC enzyme activity may provide a diagnosis as well.

Reference(s): Gyato et al., Ann Neurol 2004;; Lichter-Konicki et al., GeneReviews 2021

Q391

A man is affected by an extremely rare autosomal recessive genetic condition. What is the chance that the man’s grandchild will be a carrier of the condition?

12.5%

25%

50%

100%

Q391 answer: 50%

Q391 explanation

The man in the question is affected by a rare autosomal recessive genetic condition. Using standard terminology, we could describe his genotype as: aa (the A allele represents the wild-type allele, while the a allele harbors the pathogenic variant related to the condition). Each of his children would be expected to inherit the a allele. Since this is a rare condition, we can assume it is very unlikely that they will inherit another pathogenic variant in the gene. We can describe his childrens’ genotypes as: Aa. That is, they must all be heterozygotes, or carriers of the allele with the pathogenic variant. Each of their children (the man’s grandchildren) would have a 50% chance of inheriting the A allele and a 50% chance of inheriting the a allele, so the chance that they would be carriers would be 50%.

Reference(s): Thompson & Thompson, Genetics in Medicine, 8th ed.

ChatGPT first answer: 12.5%

ChatGPT second answer: 25%

ChatGPT explanation:

The chance that the man's grandchild will be a carrier of the condition is 25%. Autosomal recessive genetic conditions are caused by mutations in genes located on autosomes (non-sex chromosomes). A person must inherit two copies of the mutation, one from each parent, to develop the condition.

If a person inherits one copy of the mutation, they are a carrier of the condition and do not show symptoms. Since the man is affected by an extremely rare autosomal recessive genetic condition, it is likely that his spouse is also a carrier of the same condition.

When two carriers of an autosomal recessive condition have a child, the chance of the child inheriting two copies of the mutation (and thus being affected by the condition) is 25%. The chance that the child will be a carrier of the condition (and not affected) is also 25%. The chance that the child will not inherit any copies of the mutation and will not be a carrier is 50%.

Q392

You suspect that your patient has myotonic dystrophy. Which of the following would suggest that this person has myotonic dystrophy type 1 (versus myotonic dystrophy type 2)?

Arrhythmias

Cataracts

Developmental delay

Myotonia

Q392 answer: Developmental delay

<https://pubmed.ncbi.nlm.nih.gov/27858759/>

<https://www.ncbi.nlm.nih.gov/books/NBK1466/>

<https://www.ncbi.nlm.nih.gov/books/NBK1165/>

Q392 explanation

Myotonic dystrophy is an autosomal dominant condition that involves progressive myopathy and myotonia, and which can affect multiple organ systems. There are two main forms, termed myotonic dystrophy type 1, which is related to expansion of a repeat element in *DMPK*, and myotonic dystrophy type 2, which is related to repeat element expansion in *CNBP*. The conditions have overlapping clinical manifestations, though the manifestations can overall be highly variable from one patient to the next. Of note, myotonic dystrophy type 1 can involve certain features not seen in myotonic dystrophy type 2, including neonatal or early childhood weakness and respiratory insufficiency, developmental delay/neurodevelopmental disability, craniofacial manifestations, and clubfeet.

Reference(s): Meola and Cardani, J Neuromuscul Dis 2015; Schoser, GeneReviews 2020; Bird, GeneReviews 2021

ChatGPT first answer: Myotonia

ChatGPT second answer: Cataracts

ChatGPT explanation:

Myotonia would suggest that this person has myotonic dystrophy type 1. Myotonia is a symptom of both myotonic dystrophy type 1 (DM1) and myotonic dystrophy type 2 (DM2) where there is a delay in muscle relaxation after contraction. However, myotonia is more severe and more prominent in DM1 than in DM2.

DM1 is caused by an expansion of a CTG trinucleotide repeat in the DMPK gene on chromosome 19, while DM2 is caused by an expansion of a CCTG repeat in the CNBP gene on chromosome 3. Myotonia is a key symptom of DM1 and is present in all affected individuals, while it is less common in DM2, and some people with DM2 may not have myotonia.

Arrhythmias, cataracts and developmental delay are symptoms of DM1 and DM2 but not specific enough to distinguish between the two types of myotonic dystrophy, thus Myotonia is the best answer.

Q393

You follow a young adult who has been diagnosed with Lynch syndrome. Her father is also affected; he was diagnosed with colorectal cancer in his late 30s. According to current guidelines, at about what age is colonoscopy recommended for your patient?

10-15 years of age

20-25 years of age

30-35 years of age

Around 50 years of age

Q393 answer: 20-25 years of age

<https://www.ncbi.nlm.nih.gov/books/NBK1211/>

Q393 explanation

Lynch syndrome is a genetic cancer predisposition syndrome that can be caused by variants in several known genes. Affected individuals are at increased risk of colorectal cancer (CRC) as well as other cancer types. According to current guidelines, affected individuals are recommended to undergo colonoscopy with removal of precancerous polyps starting at 20-25 years of age or 2-5 years prior to the earliest CRC in the family, whichever is earlier. Colonoscopy is recommended every 1-2 years. Screening related to other cancers is also recommended for affected individuals.

Reference(s): Idos and Valle, GeneReviews 2021

Q394

A young adult has anomalies affecting the limbs, but extensive testing, exam, and history do not reveal any other medical issues, such as affecting other organ systems, or neurodevelopmental issues. What gene would most likely be involved?

BBS1

FANCA

GDF5

NIPBL

Q394 answer: GDF5

<https://pubmed.ncbi.nlm.nih.gov/16127465/>

<https://pubmed.ncbi.nlm.nih.gov/33333243/>

Q394 explanation

Pathogenic variants in many different genes can cause limb anomalies. For some genes, the associated conditions involve other organ systems as well as limb anomalies. Pathogenic variants in the gene *GDF5* can cause a variety of different conditions. Some of these conditions can involve isolated limb anomalies, such as brachydactyly or fusion of joints in the hands or feet. Other conditions related to the gene *GDF5* can involve syndromes affecting multiple organ systems. In general, identifying an anomaly should prompt careful evaluation, including to rule out other manifestations of a pleiotropic condition.

Reference(s): Seemann et al., J Clin Invest 2005; Genovesi et al., Bone 2021

Q395

You are designing a laboratory assay to test for a condition that involves a “repeat expansion” (related to a repeat disorder, like Fragile X syndrome, Huntington disease, or Myotonic dystrophy type 1). Which of the following lab techniques would most likely describe the main test methodology that would be used for this assay?

FISH testing

PCR fragment analysis

Sanger sequencing

SNP-based microarray

Q395 answer: PCR fragment analysis

<https://pubmed.ncbi.nlm.nih.gov/27540028/>

<https://pubmed.ncbi.nlm.nih.gov/29325606/>

Q395 explanation

There are multiple ways to test for repeat expansions of the type that cause conditions like  Fragile X syndrome, Huntington disease, or Myotonic dystrophy type 1, as well as many other conditions. One commonly used method is called “PCR fragment analysis.” In general, the method involves using PCR to amplify the region involved in the repeat condition. Then, by assessing the size of this amplified region, it can be possible to determine the number of repeats present. However, there are many nuances involved, and specific methods may be required depending on factors such as the number of repeats present in affected alleles. Other methods,such as involving Southern blots or sequencing-based approaches can also be used.

Kasenit et al., Clin Chem 2016; Paulson, Handb Clin Neurol 2018

Q396

A woman undergoes standard non-invasive prenatal screening (NIPS) to assess for certain genetic conditions affecting the fetus. What incidental finding would be most likely to be identified (and potentially reported) via this test?

Consanguinity

Fragile X syndrome

Maternal cancer

Tuberculosis

Q396

Maternal cancer

<https://pubmed.ncbi.nlm.nih.gov/23559449/>

<https://pubmed.ncbi.nlm.nih.gov/26168314/>

<https://pubmed.ncbi.nlm.nih.gov/30067923/>

Q396 explanation

Noninvasive prenatal screening (NIPS), sometimes called noninvasive prenatal testing (NIPT), involves examining a pregnant person’s blood sample to screen for certain genetic disorders in the fetus, such as certain aneuploidies as well as  other genetic conditions in some cases. This is possible since fetal DNA is detectable in the pregnant person’s blood. The test may also detect the presence of cancer in the pregnant person, as DNA from the cancer may also be detectable in the person’s blood.

Reference(s): Osborne et al., Prenat Diagnosis 2013; Bianchi et al., JAMA 2015; Bianchi and Chiu, New Engl J Med 2018

ChatGPT first answer: Consanguinity

ChatGPT second answer: Consanguinity

ChatGPT explanation (when asked the question and to provide an explanation, the correct answer was given):

Maternal cancer would be most likely to be identified (and potentially reported) via non-invasive prenatal screening (NIPS). NIPS is a test that analyzes cell-free DNA from the placenta, which is present in the mother's blood, to detect certain chromosomal abnormalities and genetic conditions in the fetus. The test is typically done during the first or second trimester of pregnancy.

It is also possible for NIPS to detect unexpected DNA changes that could indicate an undiagnosed cancer in the mother. Such changes can occur when cancer cells release their DNA into the bloodstream, which can then be detected in the maternal blood sample used for NIPS. This could include, but not limited to, cancers like breast, ovarian, colorectal and lung cancer.

Consanguinity, fragile X syndrome and tuberculosis are not related to NIPS test as NIPS analyzes cell-free DNA from the placenta and these conditions are not related to placenta or genetic conditions of the fetus.

Q397

Multiple United States Supreme Court cases have involved decisions that directly or indirectly involved biomedical patent questions. Which case most directly led to the current stance that human genes cannot be patented?

AMP v. Myriad

Diamond v. Chakrabarty

LabCorp v. Metabolite

Mayo v. Prometheus

Q397 answer: AMP v. Myriad

<https://pubmed.ncbi.nlm.nih.gov/27774159/>

Q397 explanation

The United States Supreme Court has addressed multiple cases related to biomedical patents, including the cases listed as options in the question. Association for Molecular Pathology v. Myriad Genetics, a 2013 Supreme Court case, resulted in the unanimous decision that naturally occurring DNA segments could not be patented (but allowed patents on cDNA).

Reference(s): Bakshi, J Law Biosci 2014; Contreras, The Genome Defense, 2021

Q398

A patient is suspected to have a genetic cause of rhabdomyolysis. A condition affecting which pathway or mechanism would be most likely?

Bile acid synthesis

Fatty acid oxidation

Peroxisomal metabolism

Urea cycle

Q398 answer: Fatty acid oxidation

<https://pubmed.ncbi.nlm.nih.gov/25678154/>

<https://pubmed.ncbi.nlm.nih.gov/25929793/>

Q398 explanation

Rhabdomyolysis refers to muscle injury and subsequent release of muscle contents into the blood. When severe, sequelae of rhabdomyolysis can include renal damage. There are many different causes of rhabdomyolysis, including non-genetic causes (eg, certain medications, toxins, or extreme exercise), as well as genetic causes. Rhabdomyolysis can be due to multiple genetic causes, including certain fatty acid oxidation disorders, glycogen storage disorders, mitochondrial conditions, and muscular dystrophies, as well as others. These conditions will often involve other signs and symptoms that can provide diagnostic clues, but rhabdomyolysis can be a clinically important feature.

Reference(s): Nance and Mammen, Muscle Nerve 2015; Scalco et al., Orphanet J Rare Dis 2015

Q399

In a person with sickle cell disease, which treatment would be instituted to help reduce vaso-occlusive sequelae, including pain episodes and acute chest syndrome?

Hydroxyurea

Penicillin

Pneumococcal vaccine

TNF inhibitors

Q399 answer: Hydroxyurea

<https://pubmed.ncbi.nlm.nih.gov/18367739/>

<https://pubmed.ncbi.nlm.nih.gov/20301551/>

Q399 explanation

Sickle cell disease (SCD) is a hematologic genetic condition related to the *HBB* gene. Overall management is multimodal related to the multiple organ systems the condition can affect. Among other clinical features, SCD involves vaso-occlusive manifestations. As part of the management of this issue, hydroxyurea can be beneficial in order to help reduce episodes of pain and acute chest syndrome.

Reference(s): Platt, N Engl J Med 2008; Bender, GeneReviews 2021

Q400

A couple has several children who appear to be affected by a genetic condition. Cytogenetic testing via peripheral blood reveals a cause in one of the parents, both of whom are healthy. Which of the following is the most likely finding in the parent?

Monosomy

Paracentric inversion

Pericentric inversion

Trisomy

Q400 answer: Pericentric inversion

Q400 explanation

There are many different types of cytogenetic anomalies; related clinical outcomes can be affected by the specific type of cytogenetic anomaly, as well as its size and chromosomal location. Pericentric inversions are chromosomal inversions that involve the centromere (the centromere is “within” the inversion). The products of meiosis for a person with a pericentric inversion can include normal gametes, balanced gametes, and unbalanced gametes. Among these, the inheritance of an unbalanced gamete can result in a recognizable phenotype in the offspring of the person with the pericentric inversion. Paracentric inversions, on the other hand, do not involve the centromere, and result in normal or balanced gametes or gametes that are not viable because they are acentric or dicentric (lack or have two centromeres). Thus, any children born to this person would be expected to have inherited either the normal or balanced gamete, and would not be clinically affected. A healthy parent would not be expected to have a monosomy or a trisomy. There are exceptions to the explanations described above, and there are also other types of cytogenetic findings (such as balanced translocations) that a person can have that can lead to medical conditions in their offspring.

Reference(s): Thompson & Thompson, Genetics in Medicine, 8th ed.

Q401

A person has a suspected genetic condition, but exome testing does not identify the cause. Subsequent genome sequencing does identify the cause, however. Based on this, what type of variant was most likely identified by genome sequencing?

Deep intronic variant

Exonic variant

Mosaic variant

Multi-gene deletion

Q401 answer: Deep intronic variant

<https://pubmed.ncbi.nlm.nih.gov/24941179/>

<https://pubmed.ncbi.nlm.nih.gov/25356966/>

<https://pubmed.ncbi.nlm.nih.gov/28837158/>

<https://pubmed.ncbi.nlm.nih.gov/31564432/>

Q401 explanation

Exome and genome sequencing can both be used to identify the causes of medical conditions. Exome sequencing focuses on the exons of known genes - the exons comprise about 1% of the genome. Most variants that cause genetic conditions appear to occur in the exome. Exome sequencing will not typically detect deep intronic changes (though such variants, if known to be disease-causing or of interest, could be included in a custom-capture kit so that it is sequenced along with the rest of the exome). Exome sequencing would be anticipated to identify exonic variants, and both exome and genome sequencing could at least theoretically identify copy number variants (CNVs) (such as deletions or duplications) that are large enough to involve multiple genes - in fact, both methods can detect much smaller CNVs. Current genome sequencing methods are less likely to be able to identify mosaicism than exome sequencing due to relative depth of sequencing, and higher depth of sequencing or different methods may be required to assess for mosaicism.

Reference(s): Biesecker and Green, New Engl J Med 2014; Retterer et al., Genet Med 2015; Stosser et al., Genet Med 2018; Kingsmore et al., Am J Hum Genet 2019

ChatGPT first answer: Deep intronic variant

ChatGPT second answer: Mosaic variant

ChatGPT explanation:

Deep intronic variant. Exome sequencing captures only a small fraction of the entire genome, typically around 1-2% of the coding regions (exons) of the genome. Deep intronic variants are variations that are located in the non-coding regions of the genome, specifically within introns, which are the regions between exons. These variants would not be captured by exome sequencing, but would be identified by genome sequencing, which captures the entire genome.

Q403

The mitochondrial genome contains multiple genes. Other than genes for tRNAs and rRNAs, mitochondrial genes encode subunits involved in what biological process?

Glycolysis

Oxidative phosphorylation

Photosynthesis

Ubiquitylation

Q403 answer: Oxidative phosphorylation

<https://pubmed.ncbi.nlm.nih.gov/11331900/>

<https://pubmed.ncbi.nlm.nih.gov/15861210/>

Q403 explanation

Oxidative phosphorylation is the final part of the primary biological process that produces ATP; both nuclear and mitochondrial genes encode subunits involved in this process. The mitochondrial genome contains 37 genes. These include 13 genes that encode subunits of the oxidative phosphorylation system, as well as 22 tRNAs and 2 rRNAs. The nuclear genome encodes more subunits of the oxidative phosphorylation system than the mitochondrial genome, as well as proteins involved in regulatory functions related to this system.

Reference(s): Smeitink et al., Nat Rev Genet 2002; Taylor and Turnbull, Nat Rev Genet 2005

Q405

At his annual check-up, a young adult mentions a strong family history of early-onset colon cancer. He undergoes colonoscopy, which identifies hundreds of polyps. Which gene is most likely involved?

APC

EPCAM

MSH6

PTEN

Q405 answer: APC

<https://www.ncbi.nlm.nih.gov/pubmed/19822006>

<https://www.ncbi.nlm.nih.gov/books/NBK1345/>

Q405 explanation

Heterozygous pathogenic variants in *APC*  result in APC-related polyposis conditions, which include familial adenomatous polyposis (FAP), attenuated FAP, and gastric adenocarcinoma and proximal polyposis of the stomach (GAPPS). Individuals with FAP can have thousands of adenomatous colonic polyps and risk of colon cancer; polyps can also occur in other parts of the GI tract, and affected individuals can have additional findings. The average age of onset of polyps in FAP is 16 years, but can occur significantly earlier or later in life.The number, age of onset, and location of polyps can differ in other APC-related conditions.

Reference(s): Half et al., Orphanet J Rare Dis 2009; Yen et al., GeneReviews 2022

Q406

Which term refers to a representation of all the different versions of genomic information within a species?

Diploid genome

Exome

Haplotype

Pangenome

Q406 answer: Pangenome

<https://pubmed.ncbi.nlm.nih.gov/33929893/>

<https://pubmed.ncbi.nlm.nih.gov/35444317/>

Q406 explanation

The pangenome refers to a reference genome that includes all genes and genetic variations for the genome of the species. Representing this pangenome can be challenging for many reasons, and there are ongoing current efforts related to representing the human pangenome.

Reference(s): Miga and Wang, Annu Rev Genomics Hum Genet 2021; Wang et al., Nature 2022

Q407

You are evaluating a patient, and suspect a certain diagnosis based on history and examination. You order exome sequencing. A diagnosis of which condition would most likely be molecularly confirmed this way?

Bardet-Biedl syndrome

Beckwith-Wiedemann syndrome

Fragile X syndrome

Prader-Willi syndrome

Q407 answer: Bardet-Biedl syndrome

<https://pubmed.ncbi.nlm.nih.gov/25356966/>

<https://www.ncbi.nlm.nih.gov/books/NBK1363/>

<https://pubmed.ncbi.nlm.nih.gov/31965078/>

Q407 explanation

There are many different types of genetic and genomic tests. Depending on the suspected condition, specific types of tests may be necessary for laboratory-based diagnosis. Bardet-Biedl syndrome can be caused by pathogenic variants affecting a number of different genes - the condition can often be molecularly diagnosed via sequencing these genes, whether via a panel, exome, or genome test (all of which can, depending on the methodologies used, detect both single nucleotide variants, as well as structural variants such as copy number variants (CNVs) and others). Before ordering a test, it can be important to check the specific methodologies involved, as the methodologies used by a given laboratory will affect which variants are detected and reported, and may vary from one laboratory to the next. The other conditions listed as choices would typically require a more specific or different assay for molecular detection, and the causative genetic changes would not usually be detected via exome sequencing.

Reference(s): Retterer et al., Genet Med 2015; Forsyth and Gunay-Aygun, GeneReviews 2020; Torene et al., Genet Med 2020

Q408

A man has bi-allelic pathogenic variants in the CFTR gene, but does not have clinical signs of cystic fibrosis. He may have congenital absence of which of the following?

Neutrophils

Pancreas

Thymus

Vas deferens

Q408 answer: Vas deferens

<https://pubmed.ncbi.nlm.nih.gov/9272157/>

<https://pubmed.ncbi.nlm.nih.gov/14551163/>

<https://www.ncbi.nlm.nih.gov/books/NBK1250/>

Q408 explanation

Bi-allelic pathogenic variants in the CFTR gene can cause cystic fibrosis (CF). Specific variants in this gene can also cause Congenital absence of the vas deferens (CAVD). There are some genotype-phenotype correlations, such that certain variants or combinations of variants are more likely to result in CAVD versus CF. CAVD involves aplasia or hypoplasia of the vas deferens and seminal vesicles, which can occur unilaterally or bilaterally, and which may come to clinical attention due to infertility. There is some clinical overlap between CF and CAVD related to respiratory as well as genitourinary manifestations.

Reference(s): Dörk et al., Hum Genet 1997; Gilljam et al., Am J Respir Crit Care Med 2004; Ong et al., GeneReviews 2017

Q409

 A pregnant person undergoes non-invasive prenatal screening (NIPS), which shows a result of 45,X for the pregnancy. According to the literature, what is the approximate positive predictive value of this result (the likelihood that this result is a true positive)?

5-7%

20-30%

50-70%

97-99%

Q409 answer: 20-30%

<https://pubmed.ncbi.nlm.nih.gov/21828326/>

<https://pubmed.ncbi.nlm.nih.gov/24657131/>

<https://pubmed.ncbi.nlm.nih.gov/25843063/>

<https://pubmed.ncbi.nlm.nih.gov/29032050/>

<https://pubmed.ncbi.nlm.nih.gov/28848190/>

<https://pubmed.ncbi.nlm.nih.gov/31123326/>

Q409 explanation

Non-invasive prenatal screening (NIPS) is a method that analyzes a blood sample from a pregnant person to identify certain conditions affecting the pregnancy. According to the primary literature, the overall current positive predictive value (PPV) of NIPS for 45,X in the fetus is estimated to be about 20-30%, with some studies providing slightly lower or higher estimates. Some sources also quote higher estimates; for example, see: <https://www.perinatalquality.org/vendors/nsgc/nipt/>. This means that about 20-30% of 45,X results from NIPS represent true positives for the fetus (of note, NIPS may also identify conditions such as 45,X mosaicism in the mother). Overall, the PPVs of certain other commonly screened autosomal aneuploidies (i.e., trisomies 13, 18, and 21) are higher than that of 45,X, while the PPVs of other conditions that are sometimes included (e.g., microdeletions) are currently lower. In the future, changes in testing methods may affect the PPV and other test characteristics.

Reference(s): Devaney et al., JAMA 2011; Porreco et al., Am J Obstet Gynecol 2014; Meck et al., Am J Obstet Gynecol 2015; Petersen et al., Am J Obstet Gynecol 2017; Bevilacqua et al., Fetal Diagn Ther 2018; Deng et al., Sci Rep 2019

ChatGPT first answer: 50-70%

ChatGPT second answer: 50-70%

ChatGPT explanation:

The approximate positive predictive value of a result of 45,X on non-invasive prenatal screening (NIPS) is 5-7%. This means that there is a 5-7% chance that the result is a true positive, and that the fetus actually has a 45,X karyotype. It is important to note that NIPS is a screening test, not a diagnostic test, and a positive result on NIPS should be confirmed with invasive diagnostic testing such as chorionic villus sampling (CVS) or amniocentesis.

Q410

A neonate has progressive encephalopathy in the first week of life. Plasma amino acids show increased levels of leucine, isoleucine, valine, and alloisoleucine. What is the most likely diagnosis?

Biotinidase deficiency

Homocystinuria

Maple syrup urine disease

Phenylketonuria

Q410 answer: Maple syrup urine disease

<https://pubmed.ncbi.nlm.nih.gov/30740402/>

<https://www.ncbi.nlm.nih.gov/books/NBK1319/>

Q410 explanation

Maple syrup urine disease (MSUD) is an autosomal recessive inborn error of metabolism caused by pathogenic variants affecting the branched-chain alpha-keto acid dehydrogenase complex. There are several different forms of MSUD, which can have different presentations and severity depending on the form. In the described scenario in the question, plasma amino acids would be expected to show increased levels of the branched-chain amino acids (leucine, isoleucine, and valine), as well as isoleucine; diagnosis can be molecularly confirmed via interrogation of the genes *BCKDHA*, *BCKDHA*, or *DBT*. Timely diagnosis is critical for optimal management.

Reference(s): Aliu et al., Ann Transl Med 2018; Strauss et al., GeneReviews 2020

Q411

Third-trimester ultrasound shows an increased (>95th %) head circumference, slightly increased abdominal circumference, and long bones below the third percentile for gestational age. What condition is most likely?

Achondroplasia

Arthogryposis

Osteopetrosis

Stickler syndrome

Q411 answer: Achondroplasia

<https://pubmed.ncbi.nlm.nih.gov/18328977/>

<https://pubmed.ncbi.nlm.nih.gov/19265753/>

<https://pubmed.ncbi.nlm.nih.gov/21105021/>

Q411 explanation

Achondroplasia, which is due to heterozygous pathogenic variants in *FGFR3*, is the most common condition resulting in disproportionate small stature. Prenatally, common features that can be observed, such as via third-trimester ultrasound (certain features will not be evident early in pregnancy) include increased head circumference, normal to increased abdominal circumference, and long bones below the third percentile for gestational age.

Reference(s): Baujat et al., Best Pract Res Clin Rheumatol 2008; Krakow et al., Genet Med 2009; Chitty et al., Ultrasound Obstet Gynecol 2011

Q412

A person has been diagnosed with Von Hippel-Lindau disease. Overall, what is the common type of neoplasm in people with this condition?

Endolymphatic sac tumor

Hemangioblastoma

Pheochromocytoma

Renal cell carcinoma

Q412 answer: Hemangioblastoma

<https://pubmed.ncbi.nlm.nih.gov/27114602/>

<https://www.ncbi.nlm.nih.gov/books/NBK1463/>

Q412 explanation

Von Hippel-Lindau disease is an autosomal dominant cancer predisposition syndrome due to pathogenic variants in the *VHL* gene. The condition involves increased risk of multiple types of benign and malignant neoplasms, including brain, spinal cord, and retinal hemangioblastoma (hemangioblastomas are overall the most common type of neoplasm in this condition - retinal hemangioblastomas are reported in up to 60% of individuals, while craniospinal hemangioblastomas are reported in up to 80% of individuals), endolymphatic sac tumors, neuoroendocrine tumors, pheochomocytoma, and renal cell carcinoma (which is also relatively common in affected people - renal cell carcinoma or renal cysts are reported in up to 75% of individuals).

Reference(s): Nielsen et al., J Clin Oncol 2016;  Leeuwaarde et al., GeneReviews 2018

ChatGPT first answer: Renal cell carcinoma

ChatGPT second answer: Hemangioblastoma

ChatGPT explanation (when asked the question and to provide an explanation, the correct answer was given):

The most common type of neoplasm in people with Von Hippel-Lindau disease is hemangioblastoma. This is a benign (non-cancerous) tumor that arises from blood vessels in the brain and spinal cord. It is also commonly found in the retina, and can cause vision problems if it grows large enough to press on the optic nerve. Hemangioblastomas are typically slow-growing and can be surgically removed or treated with radiation therapy.

Q413

A person enrolled in a research study is suspected to have a genetic condition. You use a multi-layered neural network to analyze photos of the person in order to aid the differential diagnosis. What is the best description of this type of artificial intelligence approach?

Bayesian inference

Deep learning

Diffusion model

Unsupervised learning

Q413 answer: Deep learning

<https://pubmed.ncbi.nlm.nih.gov/35145301/>

<https://pubmed.ncbi.nlm.nih.gov/35612590/>

Q413 explanation

Artificial intelligence (AI) is overall being explored and used in more and more biomedical contexts. There are many types of artificial intelligence. Machine learning (ML) is one type of AI - ML allows computers to perform tasks without explicit programming for that task. Deep learning (DL) is a subtype of ML. DL’s involves representation learning, which focuses on finding a set of features (such as a numerical vector) to represent data (such as a photograph of a person). DL is called “deep” because it uses multiple processing layers to help classify the data. For example, DL might be used to enable analysis of an image of a person’s face in order to help consider the genetic differential diagnosis for that person.

Reference(s): Hsieh et al., Nat Genet 2022; Ledgister Hanchard et al., Genet Med 2022

Q415

An infant is diagnosed with a severe form of beta-thalassemia. As part of the diagnostic work-up, what analyte would be expected to be absent or reduced on laboratory-based testing?

Factor VIII

Hemoglobin A

Hemoglobin F

Platelets

Q415 Answer: Hemoglobin A

<https://www.ncbi.nlm.nih.gov/books/NBK1426/>

<https://pubmed.ncbi.nlm.nih.gov/36125483/>

Q415 explanation

Beta-thalassemia is an autosomal recessive hemoglobinopathy caused by pathogenic variants in the *HBB* gene. The condition involves reduced production of the beta hemoglobin subunit, and has been divided into types depending on the severity; current clinical schema divide the condition into transfusion-dependent and transfusion-independent types, as well as Beta thalassemia minor/trait.. Severe forms can manifest in infancy or early childhood with microcytic anemia, jaundice, and hepatosplenomegaly. In addition to microcytic, hypochromic anemia and peripheral blood smear showing anisopoikilocytosis with nucleated red blood cells, testing will show absent or reduced hemoglobin A. Prompt diagnosis is important for management with interventions such as transfusion and chelation; gene therapy for transfusion-dependent beta-thalassemia has recently been approved.

Reference(s): Origa, GeneReviews 2021; Rubin, JAMA 2022

ChatGPT first answer: Hemoglobin F

ChatGPT second answer: Hemoglobin A

ChatGPT explanation (when asked the question and to provide an explanation, the correct answer was given):

Hemoglobin A would be expected to be reduced or absent on laboratory-based testing for a severe form of beta-thalassemia. This is because beta-thalassemia is a genetic disorder that affects the production of the beta-globin chain of hemoglobin, resulting in anemia. Hemoglobin A, which is made up of two alpha-globin chains and two beta-globin chains, will be reduced or absent in individuals with beta-thalassemia because the beta-globin chain is not being produced properly.

Q416

A person with a suspected genetic condition undergoes genomic testing, which identifies an explanatory, novel pathogenic variant in an established disease gene. The lab deposits the variant into a public database. Into which database was the variant most likely deposited?

1000 Genomes

ClinVar

GeneMatcher

gnomAD

Q416 answer: ClinVar

<https://pubmed.ncbi.nlm.nih.gov/27037489/>

<https://pubmed.ncbi.nlm.nih.gov/31777943/>

Q416 explanation

There are multiple publicly-available databases and resources that are used in genetics and genomics for both clinical and research-based applications. Among these, ClinVar (<https://www.ncbi.nlm.nih.gov/clinvar/>) is an evolving archive that includes individual, interpreted (clinically-classified) genetic variants, along with supporting evidence and related data. Many laboratories deposit variants and related evidence to resources like ClinVar in order to support the genomics community as a whole.

Reference(s): Harrison et al., Curr Protoc Hum Genet 2016; Landrum et al., Nucleic Acids Res 2020

Q418

In general (and with the understanding that there can be exceptions and atypical, rare situations), which of the following genetic changes would most likely be suspected to affect canonical splicing?

c.1845+1G>A

c.888+651T>A

c.3362A>G (p.Glu1121Gly)

c.413-24A>G

Q418 answer: c.1845+1G>A

<https://pubmed.ncbi.nlm.nih.gov/28416821/>

<https://pubmed.ncbi.nlm.nih.gov/29680930/>

<https://www.nature.com/scitable/topicpage/rna-splicing-introns-exons-and-spliceosome-12375/>

Q418 explanation

Many genetic variants that result in genetic conditions involve aberrant splicing. However, aberrant splicing can occur in different ways depending on the specific variant. One relatively common way that splicing can be affected is by variants that occur in the canonical splice acceptor and donor sites. These sites represent conserved sequences along the exon/intron boundaries. Variants that affect canonical splicing are described as affecting the “+ 1 and + 2 residues” of the  donor or the “− 1 and − 2 residues” of the acceptor splice sites. While all of the variants in the question could affect splicing (in fact, all are real examples of such variants), the first option, with the variant occurring in the “+1” residue, is most likely to affect canonical splicing.

Reference(s): Soemedi et al., Nat Genet 2017; Anna and Monika, J Appl Genet 2018

Q421

A research laboratory in the United States supports a genomic research program. The lab decides to obtain CLIA certification. This certification is most important to allow the lab to be able to do which of the following:

Bill for services

Conduct sequencing

Return individual results

Study children

Q421 answer: Return individual results

<https://pubmed.ncbi.nlm.nih.gov/18496031/>

<https://www.cms.gov/regulations-and-guidance/legislation/clia>

Q421 explanation

Oversight of clinical laboratory testing is complex, and multiple groups or agencies can be involved depending on the type of testing, the laboratory, and other factors. In the United States, the Centers for Medicare & Medicaid Services (CMS) regulates human laboratory testing through the Clinical Laboratory Improvement Amendments (CLIA). CLIA testing applies to situations where individual results are returned to individuals. There are also other types of regulatory agencies and certifications that may be involved. Regarding the other options in the question, a research-based laboratory not returning individualized results could conduct sequencing without CLIA certification. CLIA regulation is separate from the ability to bill for a test - the laboratory could return results without billing. CLIA certification is not related to the age of the individuals being tested - that would primarily be handled by the IRB (and other regulatory groups) overseeing the research program.

Reference(s): Das et al., Genet Med 2008; <https://www.cms.gov/regulations-and-guidance/legislation/clia>

Q425

Out of 100 people with pathogenic variants in a certain gene, 50 develop heart disease; out of 100 people without pathogenic variants in that gene, 10 develop heart disease. What is the relative risk of heart disease in people with pathogenic variants in that gene?

0.4

0.5

1

5

Q425 answer: 5

Q425 explanation

Relative risk can be defined as the ratio of the risk of an event in an exposure group to the risk of that event in a non-exposure group. For this question, the relative risk can be calculated as the risk (proportion) of heart disease in those with pathogenic variants to the risk of heart disease in those without pathogenic variants. This can be calculated as: (50/100)/(10/100), or 5. That is, people with pathogenic variants in the gene have five times the relative risk of heart disease as those without pathogenic variants in the gene.

Reference(s): Reference(s): Thompson & Thompson, Genetics in Medicine, 8th ed.

Q426

The wings of a bat and a bird have similar forms and functions but did not develop in this way due to a common ancestor. What is the best term for the relationship between these wing structures?

Analogous

Divergent

Embryologic

Homologous

Q426 answer: Analogous

<https://evolution.berkeley.edu/evolution-101/the-history-of-life-looking-at-the-patterns/homologies-and-analogies/#:~:text=Bird%20and%20bat%20wings%20are,the%20result%20of%20convergent%20evolution>.

Q426 explanation

Analogous structures, like bird and bat wings (specifically, their relationship as wings, not as forelimbs), have similar forms and functions due to the process of convergent evolution, but did not develop as functional wings from a common ancestor - that is, they have separate evolutionary origins as wings. In contrast, structures like the front limbs of a human and a seal or a whale are homologous, though they look very different - the structures developed from a common ancestor, but are different in form and function due to divergent evolution.

Reference(s): Thompson & Thompson, Genetics in Medicine, 8th ed.

Q427

You are analyzing whole genome sequencing data for patients with suspected  genetic conditions. For which of the following conditions would it be logical to analyze the most number of known disease-related genes to identify the molecular cause?

Cornelia de Lange syndrome

Neurofibromatosis type 1

Propionic acidemia

Tuberous sclerosis

Q427 answer: Cornelia de Lange syndrome

<https://pubmed.ncbi.nlm.nih.gov/29995837/>

<https://www.ncbi.nlm.nih.gov/books/NBK1104/>

Q427 explanation

Many genetic conditions can be caused by pathogenic variants affecting one of multiple genes - this is referred to as locus or genetic heterogeneity. Of the choices listed, Cornelia de Lange syndrome (CdLS) can be caused by pathogenic variants in the most different genes - there are at least 5-7 known genes that are known to relate to CdLS (different sources cite different numbers of genes). There are also other genetic conditions (related to other genes) that clinically resemble CdLS, such that the molecular differential diagnosis for a person with features of CdLS can be relatively broad.

Reference(s): Kline et al., Nat Rev Genet 2018; Deardorff et al., GeneReviews 2020

Q428

A teen-ager has a several year history of intellectual deterioration, ataxia, and progressive myoclonic epilepsy. His father had a condition in middle age that included ataxia, dementia, progressive psychiatric disturbances, and choreoathetosis. What condition would be most likely?

Ataxia-telangiectasia

Dentatorubral-pallidoluysian atrophy (DRPLA)

Friedreich ataxia

Huntington disease

Q428 answer: Dentatorubral-pallidoluysian atrophy (DRPLA)

<https://www.ncbi.nlm.nih.gov/books/NBK1491/>

<https://pubmed.ncbi.nlm.nih.gov/30410817/>

Q428 explanation

Dentatorubral-pallidoluysian atrophy (DRPLA), which is caused by heterozygous trinucleotide expansions in *ATN1*, is a neurologic disorder that can have different presentations according to the age of presentation (which correlates with the size of the repeat expansion). Pediatric presentations involve progressive myoclonic epilepsy, behavioral disturbances, and intellectual deterioration. Adult presentations typically involve ataxia, choreoathetosis, and dementia. In addition to different clinical manifestations, anticipation can result in marked differences in the age of presentation from one generation to the next; affected children display manifestations about 15 years prior to affected mothers and 25 years before affected fathers.

Reference(s): Veneziano and Frontali, GeneReviews 2016; Carroll et al., Tremor Other Hyperkinet Mov (NY) 2018

Q429

In general (and with the caveat that there can be exceptions), a person affected by which of the following conditions would be expected to demonstrate overt clinical manifestations at the earliest age?

Dravet syndrome

Hereditary hemochromatosis

Maturity Onset Diabetes of the Young (MODY)

Spinal and Bulbar Muscular Atrophy, X-linked 1 (Kennedy disease)

Q429 answer: Dravet syndrome

<https://pubmed.ncbi.nlm.nih.gov/28202706/>

<https://pubmed.ncbi.nlm.nih.gov/28284397/>

Q429 explanation

Many genetic conditions have congenital or very early-onset (including prenatal) clinical manifestations, though some conditions may present later in life, and there can be variable expressivity. Of the choices in the question, Dravet syndrome, a severe form of epilepsy that also involves findings such as developmental delay, hyperthermia, hypotonia, and ataxia, typically presents at the earliest age (usually with seizures), from approximately 1-18 months of age; the average age of seizure onset  is ~5 months. The most common cause of Dravet syndrome is heterozygous pathogenic variants in *SCN1A*.

Reference(s): Cetica et al., Neurology 2017; Wirrell et al., Pediatr Neurol 2017

Q430

You evaluate a young child with very convincing signs of Silver-Russell syndrome. Which test(s) would be most logical to order first?

11p15 methylation and UPD7 testing

Acylcarnitine profile

Karyotype and FISH for 22q11.2

Microarray

Q430 answer: 11p15 methylation and UPD7 testing

<https://www.ncbi.nlm.nih.gov/books/NBK1324/>

Q430 explanation

Reference(s): Saal et al., GeneReviews 2019

ChatGPT first answer: Microarray

ChatGPT second answer: 11p15 methylation and UPD7 testing

ChatGPT explanation:

The most logical test to order first for a young child with very convincing signs of Silver-Russell syndrome would be a karyotype and FISH for 22q11.2. This is because Silver-Russell syndrome is caused by a genetic alteration on the short arm of chromosome 7 (7p) or long arm of chromosome 11 (11q) in about 70% of cases. The most common genetic alteration is a loss of a segment of chromosome 11 (11q) which is known as 11q terminal deletion. The FISH test will detect this specific genetic alteration and Karyotype will confirm the presence of any structural chromosomal abnormality. It is important to note that other genetic alteration such as UPD7 and 11p15 methylation testing and Microarray are also ordered depending on the clinical presentation and FISH results.
